# Conformal prediction enables disease course prediction and allows individualized diagnostic uncertainty in multiple sclerosis

**DOI:** 10.1101/2024.03.01.24303566

**Authors:** Akshai Parakkal Sreenivasan, Aina Vaivade, Yassine Noui, Payam Emami Khoonsari, Joachim Burman, Ola Spjuth, Kim Kultima

**Affiliations:** Department of Medical Sciences, Uppsala University; Department of Biochemistry and Biophysics, National Bioinformatics Infrastructure Sweden, Science for Life Laboratory; Department of Pharmaceutical Biosciences, Uppsala University

## Abstract

Accurate assessment of progression and disease course in multiple sclerosis (MS) is vital for timely and appropriate clinical intervention. The transition from relapsing-remitting MS (RRMS) to secondary progressive MS (SPMS) is gradual and diagnosed retrospectively with a typical delay of three years. To address this diagnostic delay, we developed a predictive model that is able to distinguish between RRMS and SPMS with high accuracy, trained on data from electronic health records collected at routine hospital visits obtained from the Swedish MS Registry containing 22,748 patients with 197,227 hospital visits. To be useful within a clinical setting, we applied conformal prediction to deliver valid measures of uncertainty in predictions at the level of the individual patient. We showed that the model was theoretically and empirically valid, having the highest efficiency at a 92% confidence level, and demonstrated on an external test set that it enables effective prediction of the clinical course of a patient with individual confidence measures. We applied the model to a set of patients who transitioned from RRMS to SPMS during the cohort timeframe and showed that we can accurately predict when patients transition from RRMS to SPMS. We also identified new patients who, with high probability, are in the transition phase from RRMS to SPMS but have not yet received a clinical diagnosis. We conclude that our methodology can assist in monitoring MS disease progression and proactively identify patients undergoing transition to SPMS. An anonymized, publically accessible version of the model is available at https://msp-tracker.serve.scilifelab.se/.

## Introduction

Multiple sclerosis (MS) is an inflammatory, neurodegenerative disease affecting the central nervous system. It is a leading cause of neurological disability in young adults globally. The course of MS is heterogeneous but typically involves an early, predominantly inflammatory disease phase termed relapsing-remitting MS (RRMS) and a later, principally degenerative stage known as secondary progressive MS (SPMS). SPMS is diagnosed retrospectively, where the average delay is three years^1^. While current disease-modifying therapies are effective in RRMS, the majority have very limited benefit in SPMS, if at all. Proactive recognition of patients with progressive disease could limit exposure to ineffective medications and their side effects. Early identification of patients eventually fulfilling the criteria of SPMS would, therefore, be a valuable addition to the armamentarium of clinical practitioners, enabling meaningful intervention.

Previous studies have explored invasive and non-invasive biomarkers, including biochemical and imaging-based measures, for predicting disease progression^2,3^, and the transition to SPMS^4–11^. However, the predictive value of these markers is limited^4,12^, they lack an uncertainty measure, and they are not routinely used in clinical practice. One potential approach to timely disease progression identification is using artificial intelligence (AI) and machine learning (ML). Progress in these fields has opened up the possibility of assimilating and interpreting complex data in healthcare and is expected to be transformational^13^. Machine learning and deep learning (DL)-based methods have been developed to predict the transition from RRMS to SPMS^14^.

In a study by Manouchehrinia et al., the authors achieved an accuracy of 77 to 87% when predicting the risk of conversion to SPMS in 10, 15, and 20 years using a nomogram-based method^15^. The study used electronic health record data (EHR) from 8,825 RR onset MS patients in Sweden and was validated using 6,498 patients. However, the model was developed only using data from the first hospital visit from a certain patient, and there were no risk scores associated with each hospital visit. A similar study to predict transition to SPMS within 180, 360, or 720 days was carried out by Seccia et al, utilizing 1,624 patients with 18,574 clinical records^14^. The tool was designed to make predictions using both historical clinical records and individual follow-ups. While this study demonstrated higher specificity and recall, the precision of the predictions was lower, resulting in an increased number of false positives being included. Both studies focused solely on RRMS patients, potentially missing those who had transitioned. Additionally, the studies did not incorporate any uncertainty measure for their predictions, making the model susceptible to errors when applied to external data.

As the transition from RRMS to SPMS is gradual, with overlapping disease processes during this transitional period, developing a binary classifier is challenging^16^. More generally, the adoption of predictive AI tools in healthcare thus far has been limited by more than solely their measured performance. Significant shortcomings in the clinical setting include an inability to convey uncertainty in a given prediction^17^ and a lack of explainability or interpretability for a given prediction^18^. The explainable AI (XAI) models can help healthcare practitioners understand and more easily verify the results provided by these models.

Conformal prediction (CP) is a framework for complementing single-valued predictions from standard ML/AI classifiers with a valid measure of the prediction’s uncertainty^19^. At a specified confidence level, the conformal predictor will provide a region around the point prediction containing the true label. For instance, when predicting a patient’s RRMS or SPMS disease state, CP produces four outputs: {RRMS}, {SPMS}, {RRMS, SPMS}, and {}. If the CP output contains multiple-labels, the prediction incorporates more than one true label, thus predicting a patient to be both RRMS and SPMS. Conversely, if a CP generates empty predictions, it signifies that a valid prediction cannot be made. We have recently demonstrated that CP can substantially reduce the number of errors made by an AI classifier in grading prostate biopsies^20^, and that ML in combination with CP can aid in predicting the transition of SPMS based on biomarkers measured in cerebrospinal fluid (CSF) analysis^21^. However, this approach has not been assessed with EHR data alone, which could circumvent the need for invasive or costly biomarkers.

In this study, we develop conformal predictors for ML-assisted diagnostics in MS using clinical information from the EHR collected from 22,748 MS patients with 197,227 hospital visits. We demonstrate that the model is well-calibrated, meaning the conformal predictors are valid. This allows us to produce reliable predictive uncertainties for each patient’s hospital visit. We also show how these predictors can be used to monitor a patient’s disease progression in the spectrum between RRMS and SPMS, allowing earlier identification of patients fulfilling the criteria of SPMS. We then incorporated SHapley Additive exPlanations (SHAP) to demonstrate the contributions of clinical variables to the individual predictions and the entire test dataset^22^.

We believe this approach can assist in monitoring the disease progression, earlier identification of transition to SPMS, and provide a powerful tool for tracking interventions’ effects that can also be used in clinical trials. Finally, we have set up a publicly accessible web server deploying the ML architecture for research use only.

## Results

We trained an AI model to identify patients with diagnoses of either RRMS or SPMS using EHR data from the Swedish MS Registry (SMSReg)^23^. The SMSReg is a nationwide registry containing data from 22,748 MS patients, with 197,227 hospital visits, collected between 1972 and 2022. The registry has high validity and broad coverage, estimated to include over 80% of all people with MS in Sweden^24^. More than 850 neurologists have contributed data to the registry.

The data from the registry was processed as illustrated in Fig. 1. Only patients with an RRMS or SPMS diagnosis at the first presentation were included. Duplicate entries were removed, and individual patient records were divided into hospital visits. Fifty-six clinical parameters from the EHR were used to generate 61 derived features (Supplementary Table 1). The dataset was split into four non-overlapping subsets of patients for the training (individual patients, np=10,067), validation (np=1,078), calibration (np=2,157), and testing (np=1,080) of the models. The baseline characteristics of the patients in these four subsets were similar, as outlined in Table 1.

**Fig. 1:**
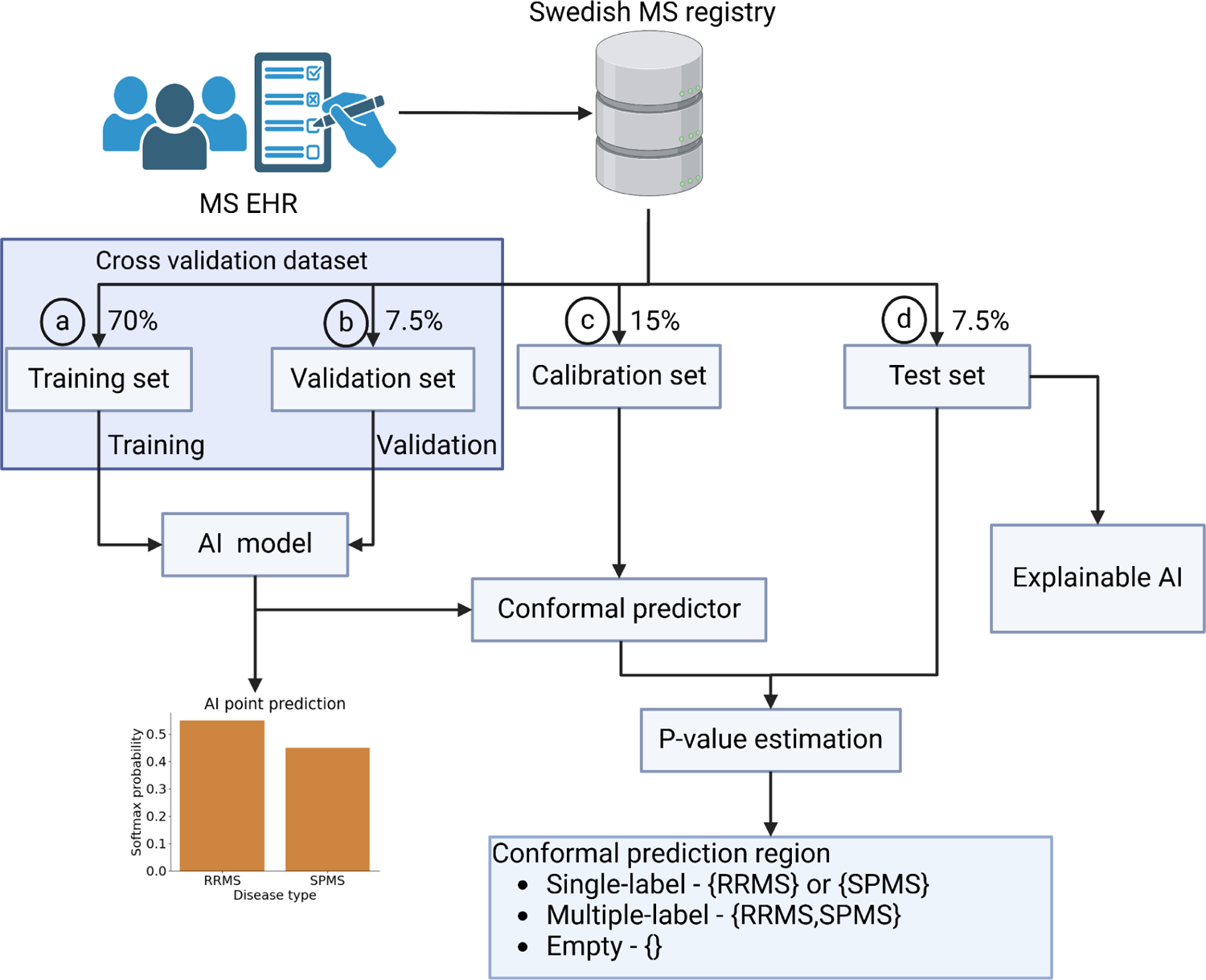
Overview of model training, validation, calibration, and testing. (a) 70% of the data was used for training, with (b) 7.5% used as a validation set for the validation. (c) 15% of the data was kept aside as the calibration set(d) 7.5% was set aside as the test set to evaluate the model efficiency.

**Table 1:** Patient characteristics in the train, valid, calibration, and test dataset. Distributions of patients with diagnoses of RRMS and SPMS. Distributions are similar in all datasets (train, valid, calibration, and test). In this table, patients recorded to have a diagnosis of RRMS at the final hospital visit were categorized as RRMS. Patients with a diagnosis of SPMS were categorized as SPMS. In the SPMS patients, patients with SPMS at initial and at final hospital visits (SPMS-SPMS) ^1^np=1,859; nv=11,320, ^2^np=193; nv=1,178, ^3^np=395; nv=2,551, ^4^np=203; nv=1,187 and patients with RRMS at initial and SPMS at final hospital visits (RRMS-SPMS) ^1^np=1,318; nv=16,171, ^2^np=158; nv=1,762, ^3^np=284; nv=3,588, ^4^np=149; nv=1,775. np=number of individual patients, nv=number of hospital visits.

|  | Train dataset<br>np=10,067; nv=79,721 |  | Valid dataset<br>np=1,078; nv=8,388 |  | Calibration dataset<br>np=2,157; nv=17,339 |  | Test dataset<br>np=1,080; nv=8,336 |  |
| --- | --- | --- | --- | --- | --- | --- | --- | --- |
| Features | RRMS<br>(np=6,890;<br>nv=52,230) | SPMS<sup>1</sup><br>(np=3,177;<br>nv=27,491) | RRMS<br>(np=727;<br>nv=5,448) | SPMS<sup>2</sup><br>(np=351;<br>nv=2,940) | RRMS<br>(np=1,478;<br>nv=11,200) | SPMS<sup>3</sup><br>(np=679;<br>nv=6,139) | RRMS<br>(np=728;<br>nv=5,374) | SPMS<sup>4</sup><br>(np=352;<br>nv=2,962) |
| Female % | 72.2 | 69.7 | 70.4 | 75.5 | 72.3 | 70.4 | 73.1 | 68.8 |
| Age ± SD | 40.4 ± 10.9 | 56.3 ± 10.8 | 39.9 ± 11.1 | 55.8 ± 10.6 | 40.5 ± 11.3 | 55.9 ± 10.8 | 40.9 ± 11.9 | 54.8 ± 10.9 |
| EDSS ± SD | 1.7 ± 1.2 | 5.6 ± 1.7 | 1.58 ± 1.1 | 5.6 ± 1.7 | 1.8 ± 1.2 | 5.6 ± 1.6 | 1.7 ± 1.2 | 5.6 ± 1.7 |
| Disease duration ± SD | 8.9 ± 7.2 | 22.1 ± 11.4 | 8.6 ± 7.9 | 21.5 ± 11.3 | 8.5 ± 6.9 | 20.9 ± 10.8 | 9.0 ± 7.4 | 21.7 ± 10.4 |
| Debut relapse age ± SD | 24.1 ± 16.6 | 26.5 ± 17.3 | 22.8 ± 16.8 | 26.5 ± 17.6 | 24.15 ± 17.0 | 28.7 ± 17.2 | 24.2 ± 17.2 | 26.0 ± 16.7 |

To account for uncertainty on an individual patient level, we used conformal prediction and assessed the model efficiency as the fraction of all the predictions, resulting in a single-label prediction. We also evaluated the model’s validity, i.e., the error rate did not exceed the pre-specified significance level of the conformal predictor, added XAI using SHAP to elucidate the features influencing the predictions, and developed a publicly available model for use in research.

### Machine learning models on EHR data produce accurate models to predict RRMS and SPMS

First, we assessed the performance of different ML models in predicting whether a patient had a diagnosis of RRMS or SPMS at a given hospital visit. Four ML models were trained: logistic regression, support vector machines (SVM), gradient-boosting (GB), random forest (RF), and a DL model (’long short-term memory’, LSTM). The latter was selected for its ability to use historical information from prior hospital visits to guide predictions for the same patient in subsequent visits.

We evaluated the ML and DL models using tenfold cross-validation on the combined training and validation datasets (individual patients, np=11,145; hospital visits, nv=88,109). Based on the macro average F1 score, the combined measure of precision and recall, the performance in discriminating between RRMS and SPMS at hospital visits was high. RF, SVM, and GB all had an F1 score of 0.91. These three models significantly outperformed logistic regression and LSTM (p< 0.05, Supplementary Fig. 1 and 2). While the three traditional ML models performed similarly to one another, RF had the lowest variation (0.905 ± 0.007), and we selected this for subsequent analysis.

In many cases, the information the different clinical variables hold is redundant. We investigated if we could identify a minimal number of clinical features used in the model without negatively impacting the overall performance. On average, the RF model performed best when we excluded the information from the patient-reported Multiple Sclerosis Impact Scale (MSIS-29), retaining 27 features (Supplementary Table S1, and Supplementary Fig. 3 and 4).

### Conformal prediction produces valid and efficient models for predicting MS diagnosis at a hospital visit

We added a valid measure of the prediction uncertainty using CP to complement the single-valued prediction from the best-performing RF model. The output p-values from the model were calibrated using the calibration dataset with data from 2,157 patients at 17,339 hospital visits. The calibration plot (Fig. 2a) demonstrates the very close correspondence between the specified significance level and the resulting observed prediction error, indicating excellent validity of the conformal predictor.

**Fig. 2:**
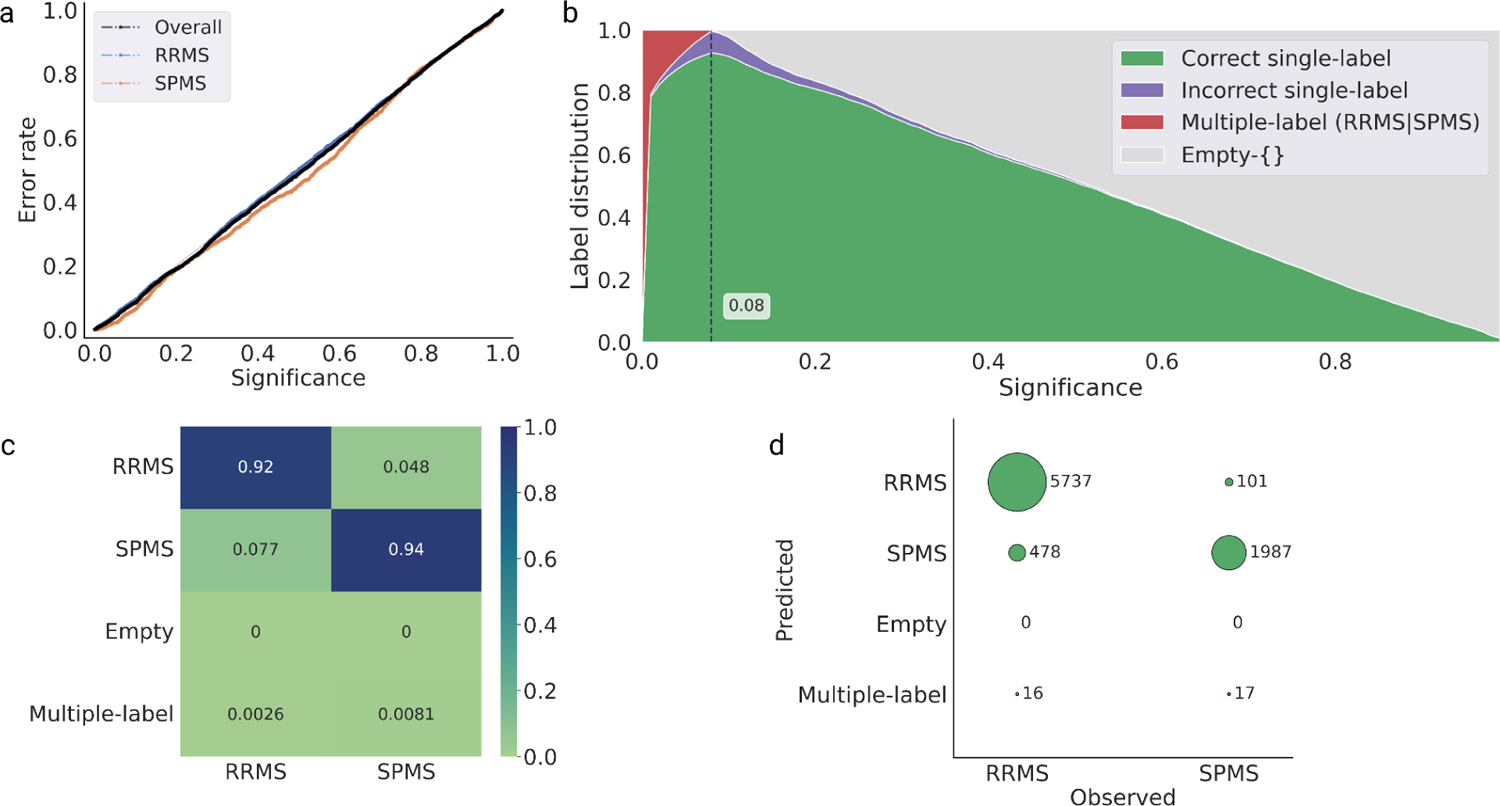
Calibration and efficiency plots on the test set data. (a) The calibration plot shows the observed prediction error on the y-axis and the prespecified significance level on the x-axis, i.e., the tolerated error rate. The observed error rate is close to the diagonal line, indicating a valid conformal predictor. (b) The efficiency plot shows the label distribution of correct single-label, incorrect single-label, multiple-label, and empty predictions for the test set at different significant levels. The plot demonstrates the expected confidence-efficiency trade-off, whereby lower significance levels (higher confidence levels) result in the conformal predictor returning an increasing proportion of multiple-label prediction and vice versa, returning an increased proportion of empty prediction at lower confidence. The confidence level corresponds to 1-significance level. The peak single-label prediction (i.e., the highest proportion of single-label predictions) is at 92% confidence, corresponding to a significance of 0.08. (c) Predictions in the test set data at 92% confidence (highest efficiency) with the predictions RRMS and SPMS indicate single-label prediction, whereas empty represents no prediction, and multiple-label represents both RRMS and SPMS prediction. (d) Normalized bubble plot showing prediction of the test set at 92% confidence.

The performance of the conformal predictor can be illustrated at different pre-specified significance levels (Fig. 2b). The conformal predictor had the highest proportion of correct single-label predictions at a significance level of 0.08, i.e., a confidence level of 92%. Consequently, we evaluated the efficiency of the conformal predictor at a 92% confidence level for predicting RRMS or SPMS in the test data set.

### Prediction with confidence for all the hospital visits using conformal prediction

The test dataset contained 1,080 patients and 8,336 hospital visits. Of these, 728 patients (5,374 hospital visits) were diagnosed with RRMS throughout, and 203 with SPMS (1,187 hospital visits). The remaining 149 patients (1,775 hospital visits) had a diagnosis of RRMS at the first visit and a diagnosis of SPMS at the last hospital visit. Based on these data, we evaluated the conformal predictor’s ability to determine the correct diagnosis i) at each hospital visit, ii) the final diagnosis for each patient, and iii) in patients with an initial diagnosis of RRMS and a final diagnosis of SPMS (“transitioning” patients). The final group was also evaluated based on the visit at which the patient was first diagnosed with SPMS relative to the conformal predictoŕs initial prediction of SPMS for each patient.

When analyzing each hospital visit, the proportion of correct single-label predictions was high (92.7%). There were a total of 579 (6.9%) incorrect single-label predictions, with no instances of empty predictions and 33 (0.4%) instances of multiple-label predictions (RRMS|SPMS) (Fig. 2c and 2d). From the incorrect predictions, in 478 cases (7.7%), the patients were erroneously predicted as having SPMS, and in 101 cases (4.8%), RRMS. Since the course of MS is heterogeneous with periods of relapses, it is not unexpected that at some hospital visits, there will be incorrect predictions. However, a closer inspection of these errors reveals that 50% of the erroneous SPMS predictions originated from only 31 patients (2.9% of all MS patients). Similarly, 50% of the incorrect RRMS predictions originated from only 11 patients (1% of all MS patients) (Supplementary Fig. 5). These results indicate that the correct single prediction efficiency of the conformal predictor is high, and the erroneous predictions often originate from a smaller fraction of all patients.

### Conformal prediction enables efficient prediction of MS diagnosis on a patient level with individual confidence measures

As SPMS is diagnosed retrospectively, we sought to evaluate the conformal predictors’ ability to predict clinical courses based on the latest available diagnosis. The overall efficiency for predicting the latest diagnosis was high (94.4%) (Table 2). There were no empty predictions, and only one patient diagnosed with SPMS received multiple-label predictions (RRMS|SPMS). At a confidence level of 92%, there were 60 erroneous predictions. Fifty-five of these (7.6%) were patients with the latest diagnosis of RRMS who were instead predicted to have SPMS. The remaining five (1.4%) patients had a final diagnosis of SPMS and were predicted to have RRMS.

**Table 2:** Prediction at final hospital visits on the test dataset with a confidence of 92% compared to the clinical diagnosis. . The predictions RRMS and SPMS indicate single-label prediction, whereas empty represents no prediction, and multiple-label represents both RRMS and SPMS prediction for the hospital visit. Np=number of individual patients. *Percentages would not add up due to rounding-off.

| Conformal prediction with a confidence of 92%. Overall efficiency was 94.4%. |  | Clinical diagnosis |  |
| --- | --- | --- | --- |
|  |  | RRMS<br>(np=728) | SPMS<br>(np=352) |
| Prediction | RRMS | 673 (92.4%) | 5 (1.4%) |
|  | SPMS | 55 (7.6%) | 346 (98.3%) |
|  | Empty-{}<br>(Empty) | 0 (0%) | 0 (0%) |
|  | Multiple-label<br>(RRMS\|SPMS) | 0 (0%) | 1 (0.3%) |

Following the prediction efficiency being markedly asymmetrical (98.3% for SPMS, 92.4% for RRMS), we investigated the conformal predictor’s output p-values for the 55 patients incorrectly predicted to have SPMS. These cases could be grouped into two categories: a majority (39 patients, 5.4%) had predictions of RRMS at the initial visit, with predictions of SPMS at later visits, while the clinical diagnosis remained RRMS (Supplementary Fig. 6).

A small group of patients (15 patients, 2.1%) persistently had predictions of SPMS at all visits despite diagnoses of RRMS, suggesting they could have SPMS already since their first presentation (Supplementary Fig. 7). One patient had conflicting predictions with p-values suggesting uncertain predictions (Supplementary Fig. 8).

### Conformal prediction coupled with XAI enables the prediction of transition states from RRMS to SPMS diagnosis

We applied our model to predict the clinical course of transitioning patients. Given the retrospective nature of the SPMS diagnosis and previously demonstrated diagnostic delays, we assessed the conformal predictor’s performance in patients who “transitioned” from a clinical course of RRMS to SPMS between the first and last visit (np=149, nv= 1,775).

Of 149 cases, the conformal predictor correctly predicted 107 (71.8%) to have RRMS at onset and later transition to SPMS (Supplementary Table 2., Fig. 3A for a patient example). In 37 cases (24.8%), the conformal predictor predicted that the patient had SPMS from disease onset. In 29 of these cases, they were predicted as having SPMS at all 213 subsequent hospital visits. The remaining eight cases had subsequent multiple-label or incorrect RRMS predictions, followed by SPMS predictions. These results display high agreement between the diagnosis and predictions; 96.6% are correctly predicted to have SPMS, and when the two deviate, the conformal predictor typically predicts the patient as having SPMS from the first presentation.

**Fig. 3:**
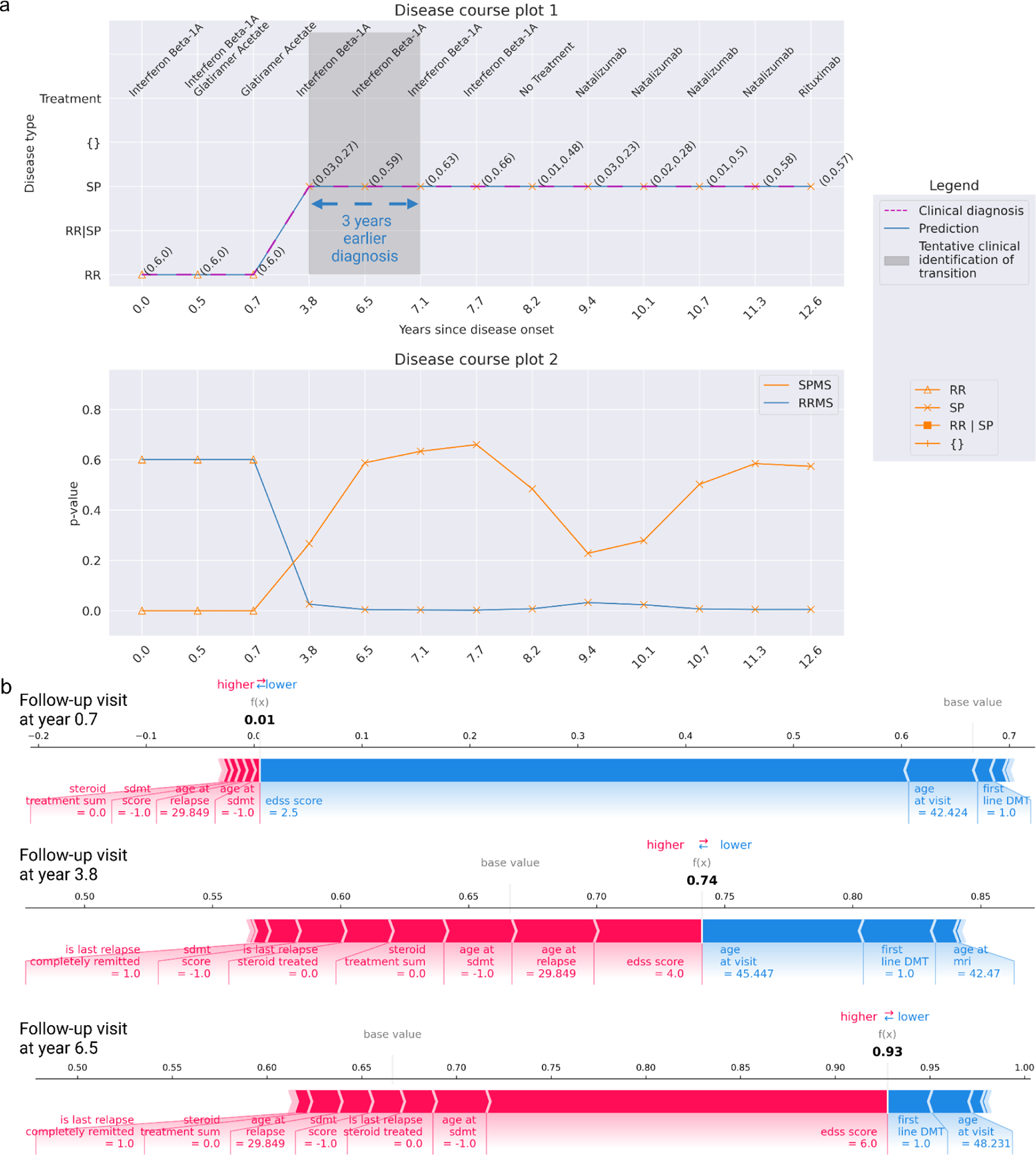
Predicted disease course at 92% confidence for an example patient with 13 hospital visits complemented with XAI. *(a)* Disease course of a transitioning patient with RRMS at the initial hospital visit and SPMS at the final hospital visit. In disease course plot 1 (top figure), the predictions consistently match the clinical diagnosis at each hospital visit, showcasing the model’s ability to identify the transition time at year 3.8. However, clinically, SPMS assessment usually transpires between years 3.8 and 7.1, as indicated by the gray zone, with an average delay of 3 years. Thus, the model identifies SPMS early, approximately 3 years in advance. Disease-modifying treatment names taken during the disease course are listed atop the figure. The disease course plot 2 (bottom figure) manifests the progression of the disease towards SPMS, indicating the disease worsening over time. A clear drop in RRMS p-value occurs between years 0.7 and 3.8, and at the same time, an increasing p-value score for SPMS is observed (between years 0.7 and 6.5). As the disability accumulates, the plot illustrates a decreasing RRMS p-value with an increasing SPMS p-value. (b) Feature contribution explanation using force plots for the predictions on the hospital visit at years 0.7, 3.8, and 6.5 of the patient. During the hospital visit year 0.7, the model predicted RRMS, driven by lower EDSS score, first-line DMT, and age at the visit. Meanwhile, the features contributing to SPMS are minimal. Conversely, in the year 3.8, the model predicted SPMS, influenced by factors such as EDSS score, age at relapse, age at SDMT, lack of steroid treatment, and complete remission of the last relapse. Features such as age at visit, first-line DMT, and age at MRI contributed towards RRMS. By year 6.5, a high EDSS score significantly influenced the prediction of SPMS, while first-line DMT and age at visit contributed to RRMS. (The results of all visits are found in Supplementary Fig. 9-11).

To aid in understanding and verifying the predictions made at each hospital visit, the weightage given to features by the model can be interpreted using SHAP (Fig. 3b). Here, we can see that conformal predictor predicts the patient to have SPMS at the hospital visit at 3.8 years, with a moderate EDSS score of 4.0, which would typically not be sufficient for an SPMS diagnosis. Almost three years later (year 6.5), the patient had a higher p-value for SPMS, which is supported by the EDSS score of 6.0.

Upon analysis of collective feature contribution using the entire test set data, the model demonstrates that EDSS and the age at the hospital visit had notable contributions compared to other features (Supplementary Fig. 12). The features SDMT score, age at debut relapse, age at MRI, first-line DMT, debut age, treatment, and age at SDMT had moderate contributions compared to the rest of the 18 features. This demonstrates the ability of CP and XAI to aid in early diagnosis, which also may assist in enabling meaningful intervention at an earlier stage.

### Predicting the timing of a change in diagnosis from RRMS to SPMS

Next, we examined the concurrence between the time (hospital visit) when the patient was diagnosed with SPMS and the prediction made by the model at 92% confidence. In 107 cases that transitioned from RRMS to SPMS (Supplementary Table 2), there was a precise time point when the conformal predictor predicted a change in disease state. In 49 cases (45.8%), the time for a change in disease state was predicted the same as the clinician has set in retrospect (Fig. 4). In 22 cases (20.6%), it was just one hospital visit in difference. In the remaining 36 cases (33.6%), the conformal predictor predicted SPMS at an earlier hospital visit (20 cases, 18.7%) or a later hospital (16 cases, 15.0%). These results display a high degree of agreement with the time for a change in diagnosis from RRMS to SPMS and the prediction made by the model. In 85% of the cases, the predictions agreed with the time for a change in diagnosis within a deviation of one visit or predicted the time for change at an earlier time.

**Fig. 4:**
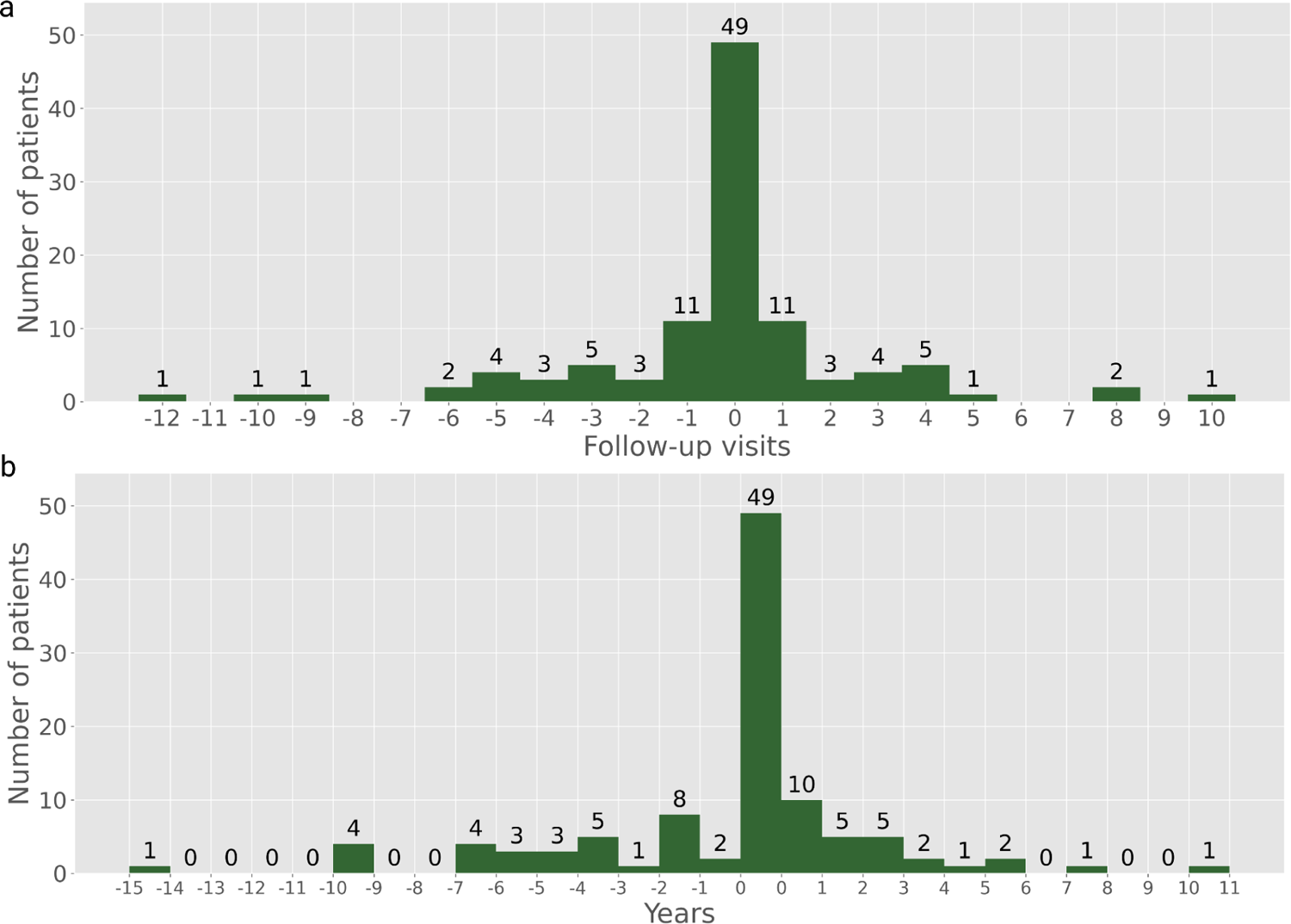
Difference of the time (hospital visits and years) for change from RRMS to SPMS diagnosis (107 patients) as found in EHR compared to the predictions at 92% confidence. An earlier prediction to SPMS is illustrated by negative values and vice-versa for a later SPMS prediction. (a) The difference in hospital visits. (b) The difference in years. Example: In Fig. 4A, 11 patients were predicted SPMS at one visit earlier and later than the clinician.

### Effects of increasing the confidence level in the conformal prediction for predicting diagnosis

In six out of the 149 cases where the patient changed their diagnosis from RRMS at debut to SPMS at the latest diagnosis, the predictions made by the conformal predictor were associated with a higher degree of uncertainty. This means that for at least two consecutive hospital visits with a period of >3 months between the visits, the patient was predicted SPMS but then changed back to RRMS, which is generally not considered possible.

Since the evaluations made of the conformal predictor were made at a confidence of 92%, where the model’s single-label predictions were highest, an alternating prediction can thus indicate that the conformal predictor cannot assign correct single predictions for these cases (Fig. 5). To investigate this further, we analyzed these cases with 95% and 99% confidence, respectively. Increasing the confidence level led to an increase in the number of hospital visits where the patient was predicted to have multiple-label (RRMS|SPMS) (Fig. 6 and Supplementary Fig. 13). By predicting multiple-label means, the model makes no errors compared to clinical diagnosis. Similar observations were found for the remaining five cases (Supplementary Fig. 13-28).

**Fig. 5:**
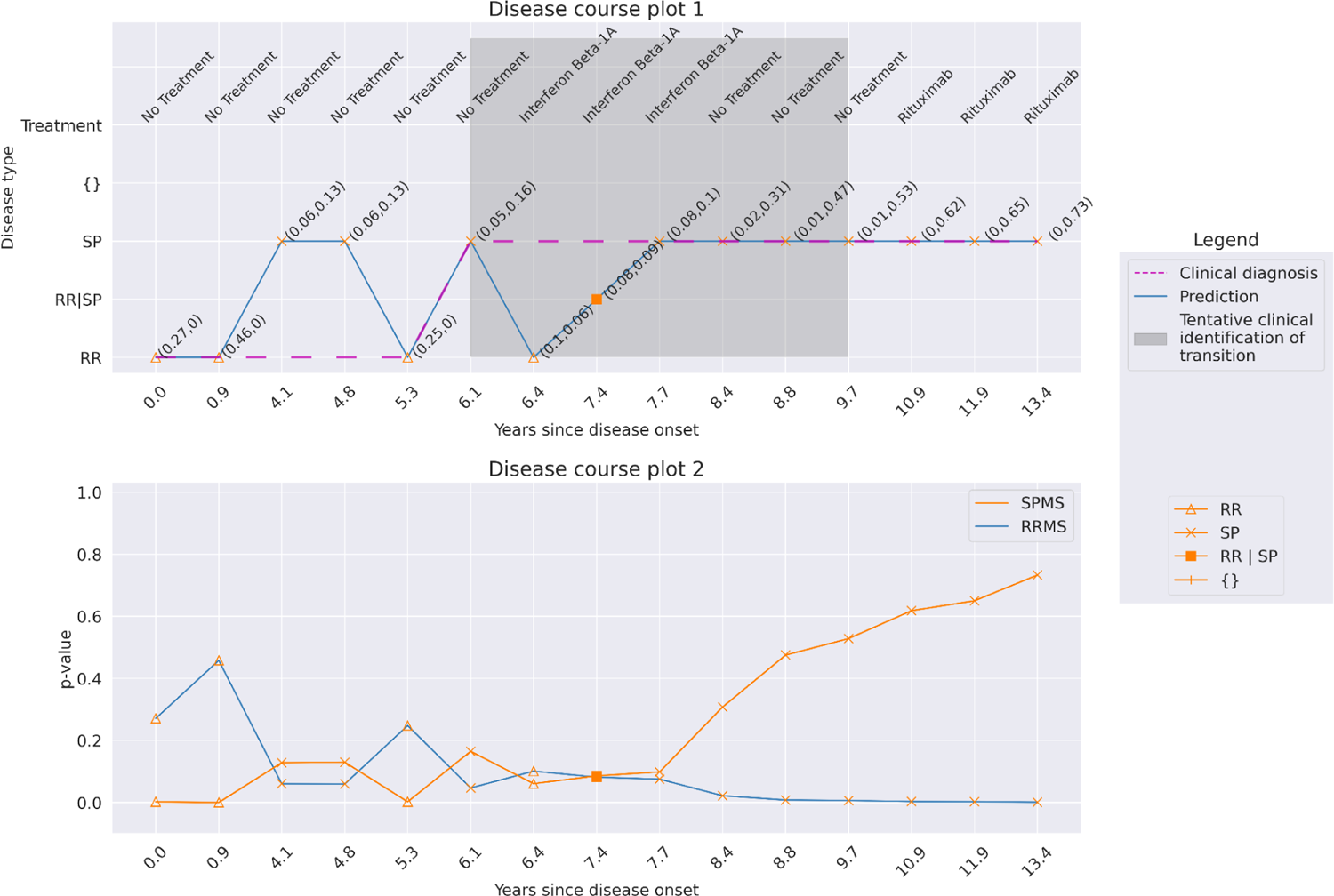
Patient 1. The prediction results at a confidence of 92% for a patient with 15 hospital visits (13.4 years). At two consecutive hospital visits, the patient was predicted SPMS with more than three months between the visits (years 4.1 and 4.8), followed by an RRMS prediction at year 5.3. This alternation is also associated with low p-values for RRMS and SPMS for the visits (disease course plot 2). Clinically, the transition occurred at the year 6.1, where the identification of the transition would tentatively be between the years 6.1 to 9.7.

**Fig. 6:**
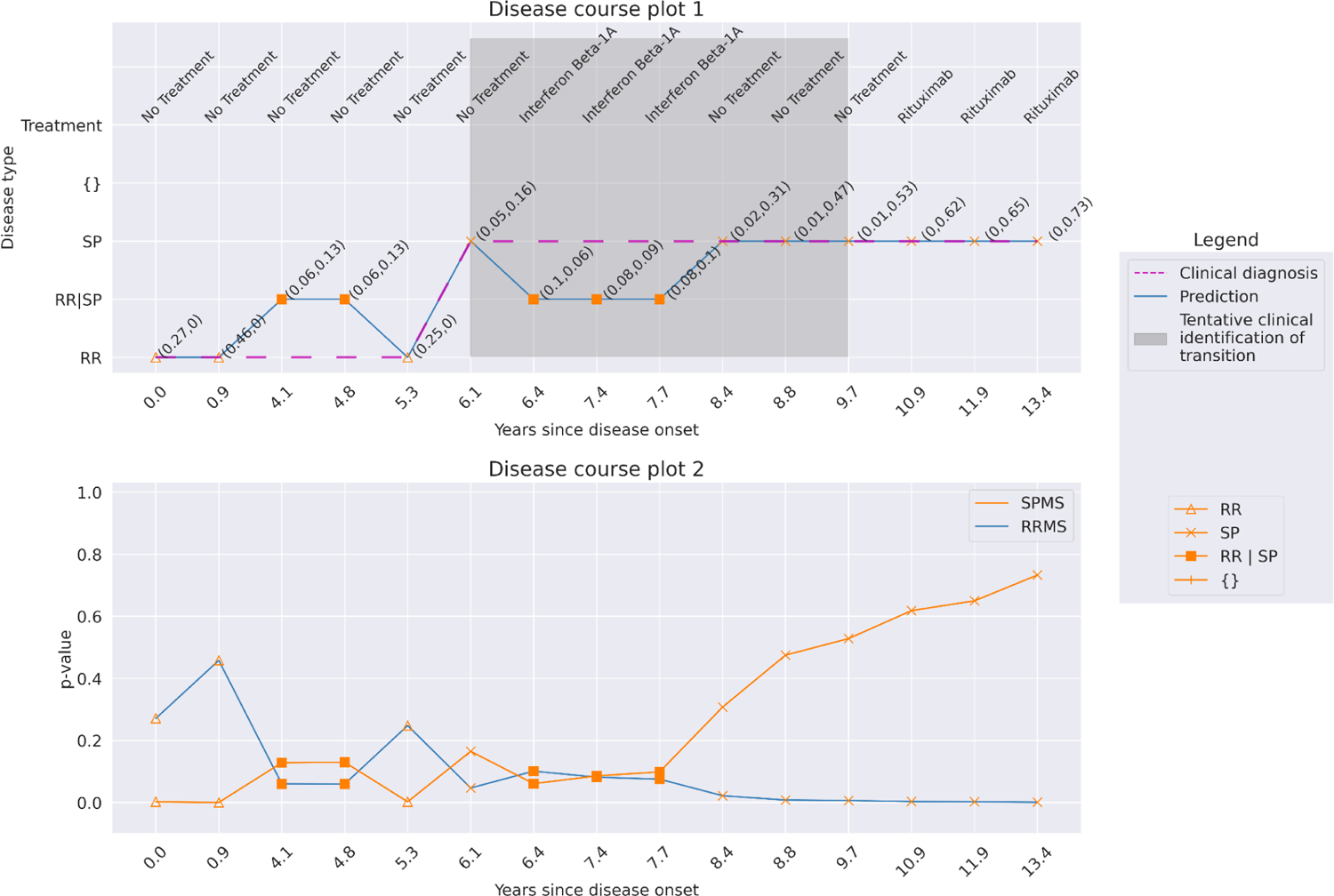
Patient 1. The prediction results at a confidence of 95% for a patient with 15 hospital visits (13.4 years). Increasing the confidence level from 92% to 95% allows for more multiple-label predictions between years 4.1 and 7.7. From year 8.4 and onwards, the model predicts only single-label SPMS. Here, the model does not make any errors but rather flags these visits 4.1, 4.8, 6.4, 7.4, and 7.7 for human analysis. The model predicts SPMS at year 6.1 with 95% confidence, aligning with the clinical diagnosis gaining earlier identification of SPMS, compared to clinical identification of SPMS between years 6.1 and 9.7.

The results of the conformal predictor for predicting diagnosis at the hospital visits when in general, increasing the confidence from 92% to 95% and 99% are found in Table 3. There are only 69 single-label misclassifications at 99% confidence (error of 0.8%), compared to 579 at 92% (error of 6.9%), but multiple-label predictions (RRMS|SPMS) increased from 33 to 1,718.

**Table 3:**
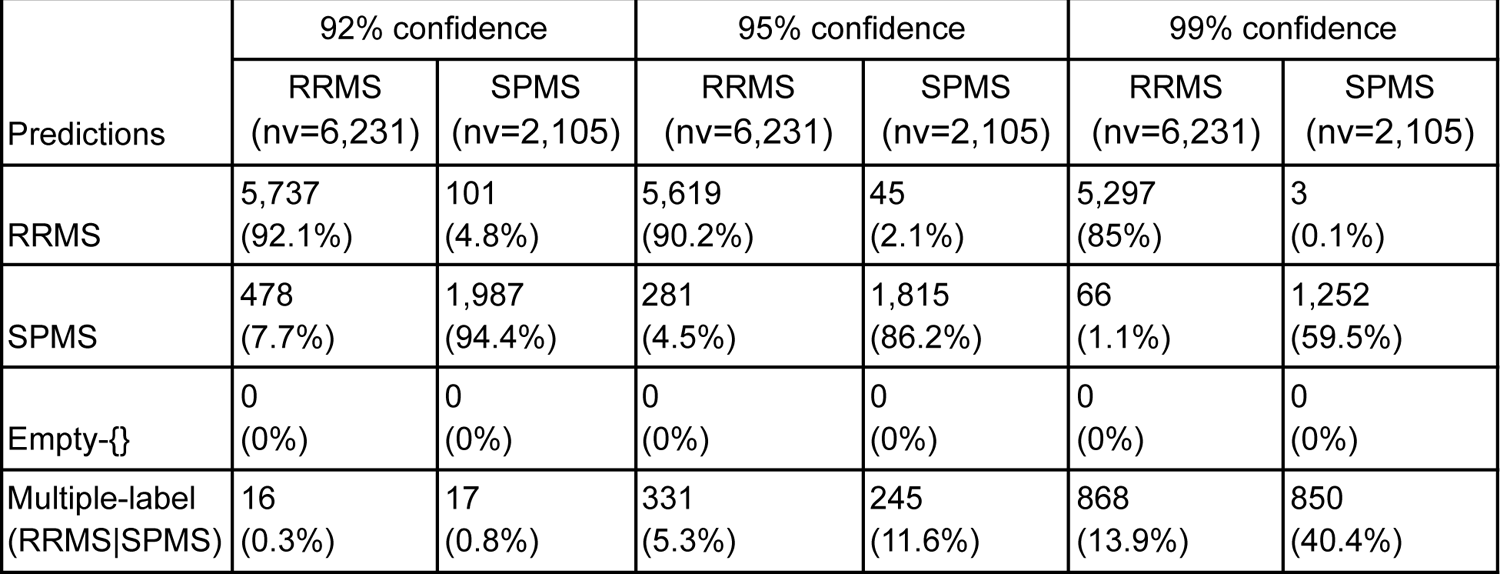
Prediction on all the hospital visits on the test dataset with a confidence of 92%, 95%, and 99% compared to the clinical diagnosis. The predictions of RRMS and SPMS indicate single-label prediction, empty represents no valid prediction, and multiple-label represents both RRMS and SPMS prediction for the hospital visit. Nv=number of hospital visits.*Percentages would not add up due to rounding off.

Since multiple-label predictions are when the model cannot distinguish between RRMS and SPMS to assign a single-label, these predictions indicate the patient is closer to or can be in transition to SPMS. We also found that some transitioning patients display non-typical disease progression, with a period of four to eight and even up to 15 years with multiple predictions, resembling an extended transition period for these cases. At a confidence level of 92%, 17 (1.6%) of all MS patients display this “atypical” disease progression, where changing the confidence level can be used to give feedback to a physician as a tool to aid in clinical decision-making.

### Publicly available *web service*

The model we have developed is based on retrospective data collected in Sweden. To aid in enabling our model to be available to other researchers, we have built a publicly available model with an interface named ‘MSP-tracker (Multiple Sclerosis Progression-tracker)’. First, there was no statistically significant difference between the performance of the *model without MSIS* model and a model that only included basic (Supplementary Table 1, Basic info) and relapse-related information (Supplementary Table 1, relapse data), ‘*Basic Info+Relapse*’ (Supplementary Fig. 3 and 4). Secondly, we removed all possibly identifiable information from the data; the year of birth was used instead of the exact date, and all other information with dates was reduced to year and month, making an anonymized model. We found no significant loss in performance difference between anonymized *MSP-tracker* and its counterpart ‘Basic Info + Relapse’ (p-value= 0.92), indicating no decay in performance by anonymizing the data.

The anonymized version of the model is available online (https://msp-tracker.serve.scilifelab.se/) and configured to accept up to 25 hospital visits. The web server can receive either direct input or from uploaded CSV files. The model can also be used at user-defined confidence levels, with output results displayed as disease course plots. The model explanation using SHAP is available post-prediction, yielding both global interpretations on the input data and individual interpretations at each hospital visit.

## Discussion

Having a clear understanding of the disease course and its current state is essential in MS, as available treatments and treatment goals vary depending on the phase of the disease. Though there are many disease-modifying treatments for RRMS, the treatments used for SPMS are few, with relatively limited efficacy^25,26^. The identification of the transition from RRMS to SPMS is made retrospectively, often with a delay of several years, and still remains a challenge^1^. Therefore, early recognition of patients with a risk of progressive disease could enable timely, meaningful interventions and also restrict unnecessary exposure to medications with associated side effects in the longer term.

We present here a first-of-its-kind predictive model that is able to distinguish between RRMS and SPMS at high accuracy, trained on data from EHR collected at routine hospital visits. To enable future usefulness in clinical settings and research, we applied conformal prediction to deliver valid measures of uncertainty in predictions on individual patient levels. We successfully produced a theoretical and empirically valid model with the highest efficiency at 92% confidence level and demonstrated on an external test set that it enables effective prediction of a patient’s clinical course with individual confidence measures (Fig. 7).

**Fig. 7:**
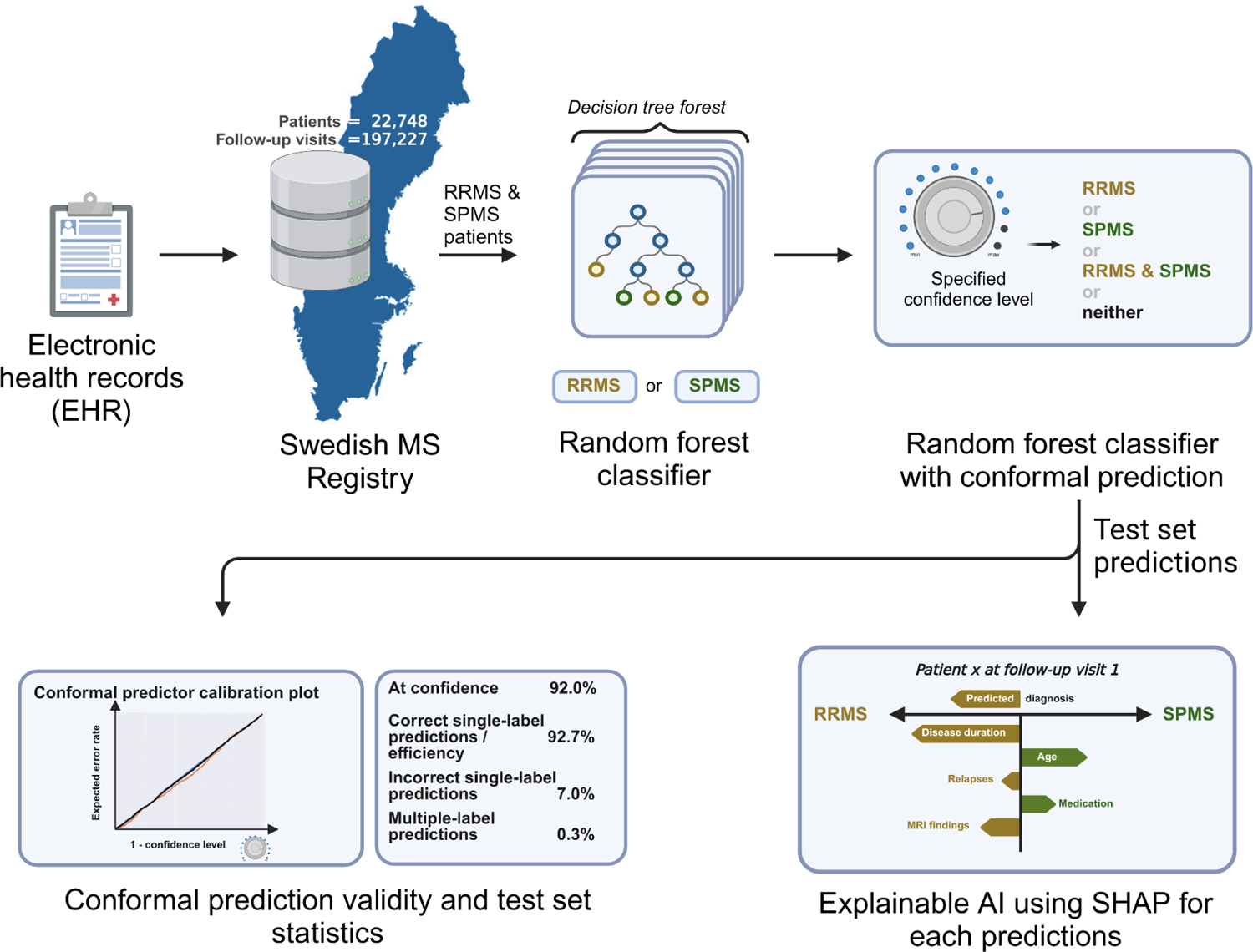
Summary figure illustrating a comprehensive view of the entire project. A predictive model, capable of distinguishing between RRMS and SPMS, was trained on EHR data collected during hospital visits in Sweden. Conformal prediction was incorporated to provide valid measures of uncertainty for predictions at the individual patient level. The model showed proof of validity and exhibited high performance on the test set data. Additionally, SHAP was utilized to understand the contribution of features in each prediction.

Clinical AI tools must convey predictive uncertainty for each individual patient^17^. We have recently demonstrated that CP-enabled AI can support predictions with user-defined confidence^20^. A well-performing CP should ideally generate fewer unreliable predictions. In this study, the model delivers 78% single-label predictions and 20% unreliable predictions at 99% confidence, marking the error rate below 1%. When faced with unreliable predictions, the explainable AI becomes essential to elucidate the reason behind these predictions. Both CP and explainable AI equip tools for users to scrutinize and analyze the occasionally unreliable predictions. Here, unreliable predictions are identified, thus allowing deeper analysis by an expert neurologist The clinical course for the transition from RRMS to SPMS has been defined by Lublin et al^27^ as the “progressive accumulation of disability after [an] initial relapsing course” assessed, at a minimum, annually^28^. As no “gold standard” criteria exist beyond this^27,29^, the SPMS diagnosis ultimately rests on the individual clinician’s judgment, primarily using the patient’s history and the clinical examination. Lorscheider et al.^30^ have since further developed the definition of SPMS by assessing the performance of standardized criteria against the “ground truth” of a consensus diagnosis by independent, expert neurologists. Multiple permutations of seven different EDSS-related criteria were generated and tested against the independent, consensus diagnosis with the best-performing permutation proposed as standardized criteria for SPMS, demonstrating performance comparable to the physician’s diagnosis.

While not widely used in clinical practice, their standardized criteria did show a notably higher sensitivity than the physician diagnosis and identified SPMS patients around three years earlier than the physician, though the specificity was lower. Using these criteria, it has been demonstrated that at the time of SPMS diagnosis, individuals will typically have EDSS scores above 4^29,30^ with disease durations greater than ten years^30^. The importance of the EDSS score in determining the transition from RRMS to SPMS is also reflected in our model, which had the largest contribution to predictions. On the other hand, other factors, such as the age at visit or time since diagnosis, also contributed substantially to the predictions in our model, but were not considered as part of the criteria described by Lorscheider et al. More importantly, the now proposed approach enables objective predictions, trained, validated, and calibrated on more than 13,000 patients with over 105,000 hospital visits. Each prediction is complemented with a measure of uncertainty, and the CP enables tracking the disease progression over time.

Multiple approaches have been described to predict the conversion from RRMS to SPMS^10,14,15,31^. In the study by Manouchehrinia et al., data from multiple cohorts was utilized, and the prediction method was specifically designed for patients with RRMS^15^. However, the model may not be applicable to patients initially diagnosed with SPMS, a limitation shared with studies by Seccia et al^14^ and Skoog et al^10^. In contrast, Ziemssen et al. categorize patients as RRMS, SPMS, or transitioning, using EHR and questionnaire data addressing this limitation^31^. In comparison, our study utilized and exhibited superior results and implemented an uncertainty measure for each hospital visit of a patient. Importantly, our model does not rely on additional questionnaire data and was able to demonstrate the progression of the disease for individual patients, which sets it apart from previous approaches. Moreover, the other models are susceptible to systematic differences between the training and external data or data drift over time. Using conformal prediction provides a robust means of handling uncertainties and addressing potential shifts in data over time^20^.

Conformal prediction proves valuable in recognizing new data that deviates from the characteristics of the training data. This is relevant when predicting an external dataset or when encountering data that the model has not seen before. This study has trained the model on data from over 850 clinicians and more than 60 Swedish hospitals. Although the model exhibits notable performance, this might not hold true when predicting external data or on unfamiliar cases within Sweden, signifying the use of CP to identify these. More than 80% of all Swedish MS patients are present in the Swedish MS registry, and data from other countries has not been part of this study, so generalization at large poses a challenge that needs to be addressed. Data drift over the years can also lead to an increase in unreliable predictions, which warns the recalibration necessary for the underlying AI and CP system.

The major strength of this study is using CP for predicting disease transition and disease state at each visit, thereby outlining the disease course of a patient. By basing predictions on clinical data already collected during hospital visits, the need for additional data collection, such as biological markers or questionnaires, is eliminated, thereby facilitating easier implementation and integration of the model in healthcare and research settings. Moreover, the model was also integrated with explainable AI, facilitating easier interpretation and assignment of labels for predictions regarded as unreliable.

The limitation of this study is the absence of analysis of the prospective data collected from the clinics. By conducting prospective data analysis, the practicality of the model at the clinics can be evaluated. Moreover, the model has not undergone evaluation using external data from other cohorts outside of Sweden. Validating CP on external data could show the potential of the model. To aid in this process, an anonymized version of the model is available online (https://msp-tracker.serve.scilifelab.se/).

## Methods

### Ethical approval

The study was approved by the Ethics Review Authority at Uppsala University (Dnr 2021-00702).

### Dataset and Quality Control

The data was obtained from SMSreg^23^, containing 22,748 patients with 197,227 hospital visits, with clinical measurements collected during each hospital visit, collected between 1972 and 2022. The data was cleaned for duplicates and missing essential data points for Expanded Disability Status Scale (EDSS) score, date of birth, progress during each visit such as RRMS/SPMS/PPMS, and debut date. The data comprises patients with transition (initial diagnosis as RRMS and final diagnosis as SPMS), RRMS (initial and final diagnosis as RRMS), and SPMS (initial and final diagnosis as SPMS). For the RRMS patients, all the visits within two years of the last visit were removed as their clinical endpoint had not yet been determined. This ensures the removal of all the unidentified transitions from the data. After quality control and removal of PPMS patients, 17,045 patients with 143,053 hospital visits were retained.

A hospital visit consisted of age and EDSS measured at the visit. For each visit, the last collected clinical measures such as treatment, clinical assessment tests, relapse data, MRI data, and MSIS data (supplement table S1) were appended, along with the age at which these measures were collected. For therapeutics, the drugs/treatments were categorized into first-line, second-line DMT, relapse treatment drugs, stem cell treatment, and any other drugs (supplement table S1).

For data relating to relapses, the total number of occurrences of different categories of relapses (including unilateral optic neuritis, sensory/afferent monofocal relapse, multifocal relapse, and relapses requiring steroid treatment) were summed up until the day of the hospital visit before appending. Additional information regarding treatment received for the relapse and remission of the last relapse was also included as binary variables. For the MRI data, the number of T2-weighted lesions, and the number and site of T1-weighted gadolinium-enhancing lesions (ie. brain vs. spinal cord) were considered. Each type of lesion was binned into three groups based on the number of lesions present at the hospital visit: 1) ≤9, 2) >9 and ≤20, 3) >20 lesions.

For a patient, during the initial hospital visit EDSS, age at visit, age at diagnosis, age at debut relapse and sex were recorded. However, certain parameters may be missing as they have not yet been measured. The missing values, including those for other parameters of the patient, were imputed using the value −1.

### Data splitting

There were three possible types of data for a patient: a patient having hospital visits with only RRMS, only SPMS, or RRMS at debut, and SPMS at the latest. To maintain an even distribution of these patients across the data splits, a stratified split was applied, grouping all the visits associated with a patient in the same split of the data. Thereby dividing the data into four subsets: training (70%), validation (7.5%), calibration (15%), and test (7.5%) datasets (Table 4). The validation set is created to optimize the deep learning model, and therefore, for traditional machine learning models, the validation dataset is merged with the training set for training. For cross-validation used in this study, the training and validation sets are merged for both deep learning and machine learning models and used for training.

**Table 4:**
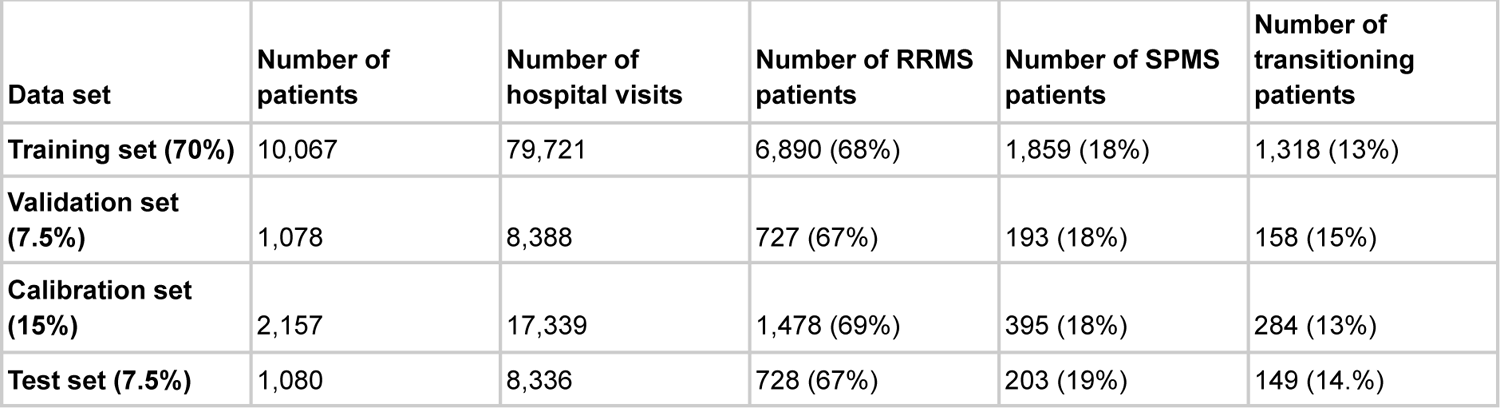
Data splits created from SMSREG data. . The dataset is divided into train, valid, calibration, and test sets, each containing unique patients and their EHR.

| Data set | Number of patients | Number of hospital visits | Number of RRMS patients | Number of SPMS patients | Number of transitioning patients |
| --- | --- | --- | --- | --- | --- |
| Training set (70%) | 10,067 | 79,721 | 6,890 (68%) | 1,859 (18%) | 1,318 (13%) |
| Validation set (7.5%) | 1,078 | 8,388 | 727 (67%) | 193 (18%) | 158 (15%) |
| Calibration set (15%) | 2,157 | 17,339 | 1,478 (69%) | 395 (18%) | 284 (13%) |
| Test set (7.5%) | 1,080 | 8,336 | 728 (67%) | 203 (19%) | 149 (14.%) |

### Architectures

Five methods were employed for prediction, 1) logistic regression (LR) 2) support vector machines (SVM) 3) gradient-boosting (GB) 4) random forest (RF) and 5) DL model using long short-term memory (LSTM). For SVM, GB and RF, gridsearch CV was carried-out to obtain the best performing model. For SVM, the RBF kernel was utilized with the parameter gamma set to 0.0001 and the regularization parameter C set to one. For GB, number of estimators was 50, a minimum sample split of two with criterion set to friedman_mse with exponential loss. For the RF architecture, minimum samples per leaf were five with the criterion as gini and the number of estimators was set to 150. The deep learning model was an LSTM with one layer having a hidden cell size of 256, combined with an fully connected multi-layer perceptron with two hidden layers and an output size 64.

### Conformal prediction

Conformal prediction (CP) is a framework built on top of any ML model to retain the error rate of the prediction to a pre-specified level^19^. CP is model agnostic (meaning, it can be implemented on all models) and is implemented on top of a prediction algorithm. Unlike single-valued output from a prediction algorithm, CP produces a prediction region containing a set of class labels for classification and a confidence interval for regression. Using CP, a non-conformity measure α_i_ is calculated for an object *i* using a non-conformity function *h(x_i_)*, where *x* represents the features and *h* represents a scoring algorithm such as a machine learning algorithm. When applied to a classification problem, at first, non-conformity α_i_ is calculated for all the n instances in the calibration set, yielding α_1_, …, α_n_. During the prediction phase, the non-conformity α_n+1_ from a test instance is used to calculate a set of p-values for each class label using equation 1, which ranks the α_n+1_ against all the α_1_, …, α_n_. Using a statistical test and employing a confidence cutoff (1-significance), such as 95%, implying a significance of 0.05, all the labels with a p-value greater than or equal to 0.05 are included in the output prediction, resulting in single-label, multiple-label, or empty predictions.

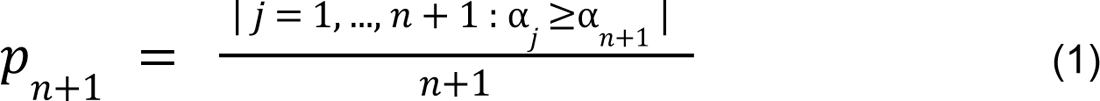

For a binary classification, the possible output from CP are {0}, {1}, {0,1}, or {}. A smaller prediction region with only a single-label ({0} and {1}) is more desired and efficient, for explaining the output of the model. A multiple-label prediction ({0,1}) is when multiple class labels have higher confidence, and the model is unable to determine between the two class labels. This occurs when uncertainty arises in assigning a single-label for the prediction. Although these predictions can be harder to interpret, they are not incorrect *per se*. These can be interpreted as unreliable/uncertain predictions, and can be flagged for manual or expert inspection to determine the correct class label. The empty set ({}) predictions are obtained if the confidence is low on both class labels, and it occurs when the input data differs from the data the model is trained on. This can highlight systematic differences between training and external data or data drifts that happened over time.

The desired CP confidence can be set by the user during the prediction time. At higher confidence, the probability of having the correct label in the output prediction set increases, yielding a wider prediction region (increase in multiple-label predictions). Likewise, lower confidence produces a smaller prediction region (increase in single-label predictions and empty predictions).

There are two ways of calibrating a conformal predictor: 1) transductive framework and 2) inductive framework. In transductive CP, for each new instance during the predictive phase, all the data is used to calculate the conformity score, making it necessary to retrain the model for every datapoint in the calibration and test set. Though this method is more robust to outliers and anomalies in the dataset, the computational demand makes it unusable for large datasets and deep learning algorithms. Inductive CP (ICP), on the other hand, is built using training and calibration datasets and is applied on the test dataset. The calibration dataset is identically independently distributed (IID) data from the training dataset. The lower computational demand and easiness of recalibrating the model make ICP popular in many fields.

In this project, we use the ICP framework, by using 15% of the available data as a calibration dataset. The basic implementation of CP considers the error rate on a population level. Making the error rate on one label to be lower than the other label. To overcome this, Mondrian CP was used to achieve a predefined error rate within each class label. Instead of tuning on the entire population, the CP was tuned on each class label. This enables reliability in prediction on an individual level, making the model applicable for clinical use, as we are more interested in individual predictions than population-level prediction in a clinical setting.

### SHAP

SHapley Additive exPlanations (SHAP) uses a game theoretic approach to generate explainable and interpretable output from a machine learning model^22^. Using this framework, SHAP values can be calculated for each feature in a data point by contrasting predictions with and without the presence of a specific feature. This process is iteratively applied for the entire dataset, resulting in the generation of SHAP values for each feature across the dataset. The difference in the impact of the feature for a prediction reveals positive or negative contributions for both individual predictions and the prediction on the entire dataset. Thus, SHAP allows us to calculate both the global interpretation, giving insight into the overall importance of features in the dataset, and also for the individual predictions, interpreting the rationale behind the output using feature contribution.

Providing explanations for individual predictions holds significant importance within clinical settings. This offers a better understanding and increases the reliability of the predictions^32^. In this project, we explain each prediction using force plots. In these plots, SHAP values for individual features are plotted along the x-axis, where each feature is represented by a bar, with the length of the bar corresponding to the magnitude of the feature’s impact and the colors indicating positive values in red and negative values in blue. The visual nature of these plots helps us understand why the model made a specific prediction.

In contrast, the global understanding of the model is achieved using a summary plot and a beeswarm plot. Both these plots provide a comprehensive overview of the importance of features in the entire dataset. The y-axis displays features ranked according to their importance, with features having a higher impact on the predictions at the top. The x-axis in the summary plot represents mean absolute SHAP values, displaying the global importance of the features. In the beeswarm plot, the x-axis represents the SHAP values and their importance, color-coded according to feature value. The SHAP value of a feature from each data point is plotted, with overlapping SHAP values jittered in the y-axis to accommodate and form a distribution.

## Data availability

The data used in the study cannot be shared to protect the privacy of individuals. All the data can be obtained by applying through SMSreg. All the necessary codes used are given in the GitHub repository https://github.com/caramba-uu/MSP-tracker.git.

## Code availability

The code used for data pre-processing, the final model, and the web server are available at https://github.com/caramba-uu/MSP-tracker.git.

## Acknowledgments

The authors like to acknowledge SciLife serve for hosting the online version of the tool MSP-tracker.

## Author contributions

The project was conceptualized, and results were interpreted by KK, JB, OS, and APS. The methods were developed by KK, OS, JB, and APS. APS implemented the methods and analysis. APS and PE were involved in reviewing the methods. APS, KK, and YN wrote the manuscript, and all the authors were involved in the review and editing process. All authors read and approved the final manuscript.

## Supplementary Information

**Supplementary Table 1:** All the derived features from the clinical features used for training of the model. The data contained missing values for clinical assessment test (60-73%), relapse data (12%), MRI data (26%) and MSIS-29 (59%). These missing values were imputed to form the complete dataset.

| Feature | Feature type |
| --- | --- |
| **Visit information (Basic info)** |  |
| **Diagnosis age** | Approximate age at which the disease was diagnosed |
| **EDSS score** | EDSS score measured during the visit |
| **Age at visit** | Age of the patient at the visit when the EDSS score was given |
| **Patient information** |  |
| **Sex label** | Sex of the patient |
| **Drug treatment** |  |
| **No treatment** | Whether the patient has not received any treatment during this visit |
| **First-line DMT** | Whether the patient is undergoing first line (disease-modifying therapies) DMT |
| **Second-line DMT** | Whether the patient is undergoing second-line DMT |
| **Other drugs** | Whether the patient is consuming any other drugs/treatment |
| **Relapse treatment drugs** | Whether the patient is receiving drugs for relapse |
| **Stem cell treatment** | Whether the patient has received hematopoietic stem cell transplantation |
| **Clinical assessment tests** |  |
| **EQ5D score** | EQ5D score during the visit |
| **Age at EQ5D** | Age at EQ5D was given |
| **SDMT score** | Symbol Digit Modalities Test (SDMT) score during the visit |
| **Age at SDMT** | Age at SDMT was given |
| **Relapse data** |  |
| **Mono on sum** | Sum of unilateral optic neuritis relapses |
| **Monofocal sum** | Sum of other monofocal relapse |
| **Multi focal sum** | Sum of multifocal relapse |
| **Afferent non on sum** | Sum of sensory/afferent non-optic neuritis relapses |
| **Steroid treatment sum** | Sum of steroid treatments received |
| **Is last relapse steroid treated** | If the last relapse was treated with steroids |
| **Is last relapse completely remitted** | Was the last relapse completely remitted |
| **Age at debut relapse** | Age at first relapse |
| **Age at relapse** | Age at relapse |
| **MRI data** |  |
| **T2 lesion category** | Number of T2-weighted MRI lesions (binned) |
| **Brain barrier lesion category** | Number of T1-weighted gadolinium-enhancing brain lesions (binned) |
| **Spinal barrier lesion category** | Number of T1-weighted gadolinium-enhancing spinal cord lesions (binned) |
| **Age at MRI** | Age at the time that the MRI was performed |
| **MSIS data** |  |
| **MSIS 01 - MSIS 29**<br>**(29 questions)** | Hobart et al<sup>1</sup> |
| **MSIS physically** |  |
| **MSIS psychologically** |  |
| **MSIS physically 100** |  |
| **MSIS psychologically 100** |  |
| **Age at MSIS** | Age at the time that the MSIS was administered |

**Supplementary Fig. 1:**
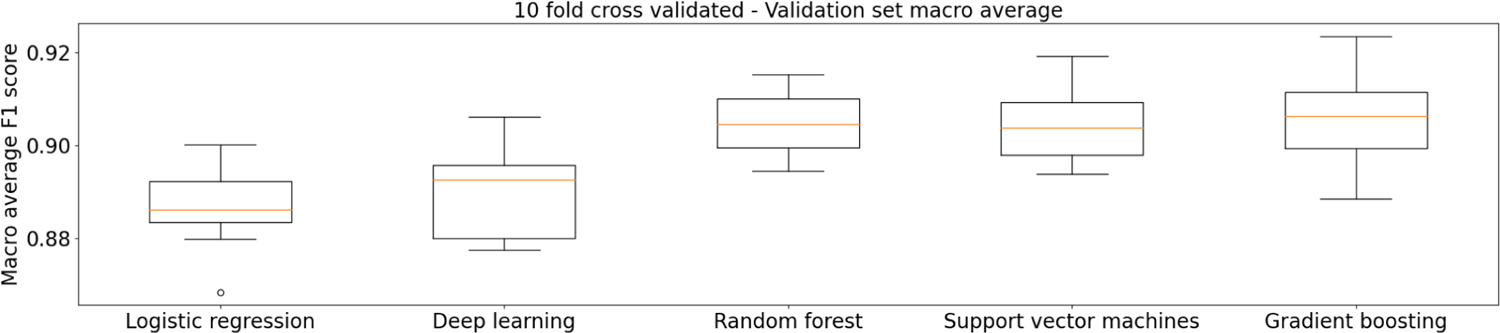
Box plot showing F1 scores from the 10-fold CV of traditional machine learning models and deep learning model. The random forest, support vector machines, and gradient boosting had better performance when compared to logistic regression and deep learning models. All the models showed an F1 score of >0.88.

**Supplementary Fig. 2:**
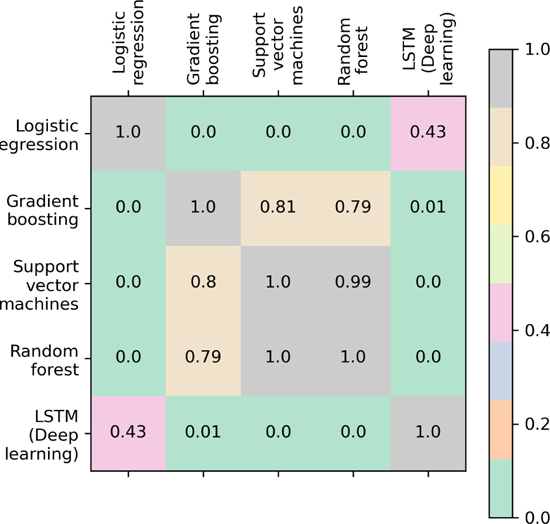
Randomization test (p-value<0.05) on F1 scores from 10-fold CV of traditional machine learning models and deep learning models. Significant p-values represent a higher or lower performance between the models. Comparing the box plot supplementary Fig. 1, random forest, support vector machines, and gradient boosting models performed significantly better than both the deep learning model and logistic regression.

**Supplementary Fig. 3:**
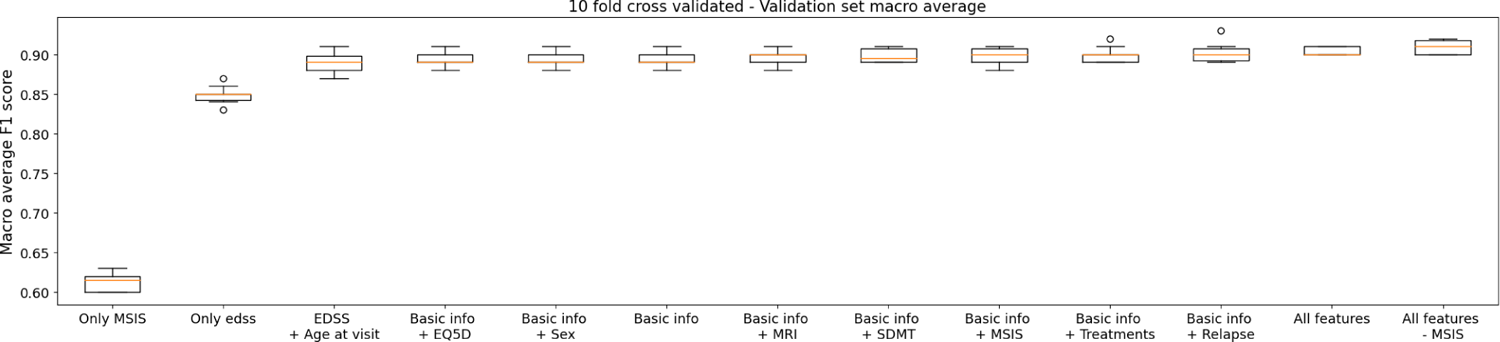
Box plot showing F1 scores from the 10 fold CV of RF models trained on combination of features.

**Supplementary Fig. 4:**
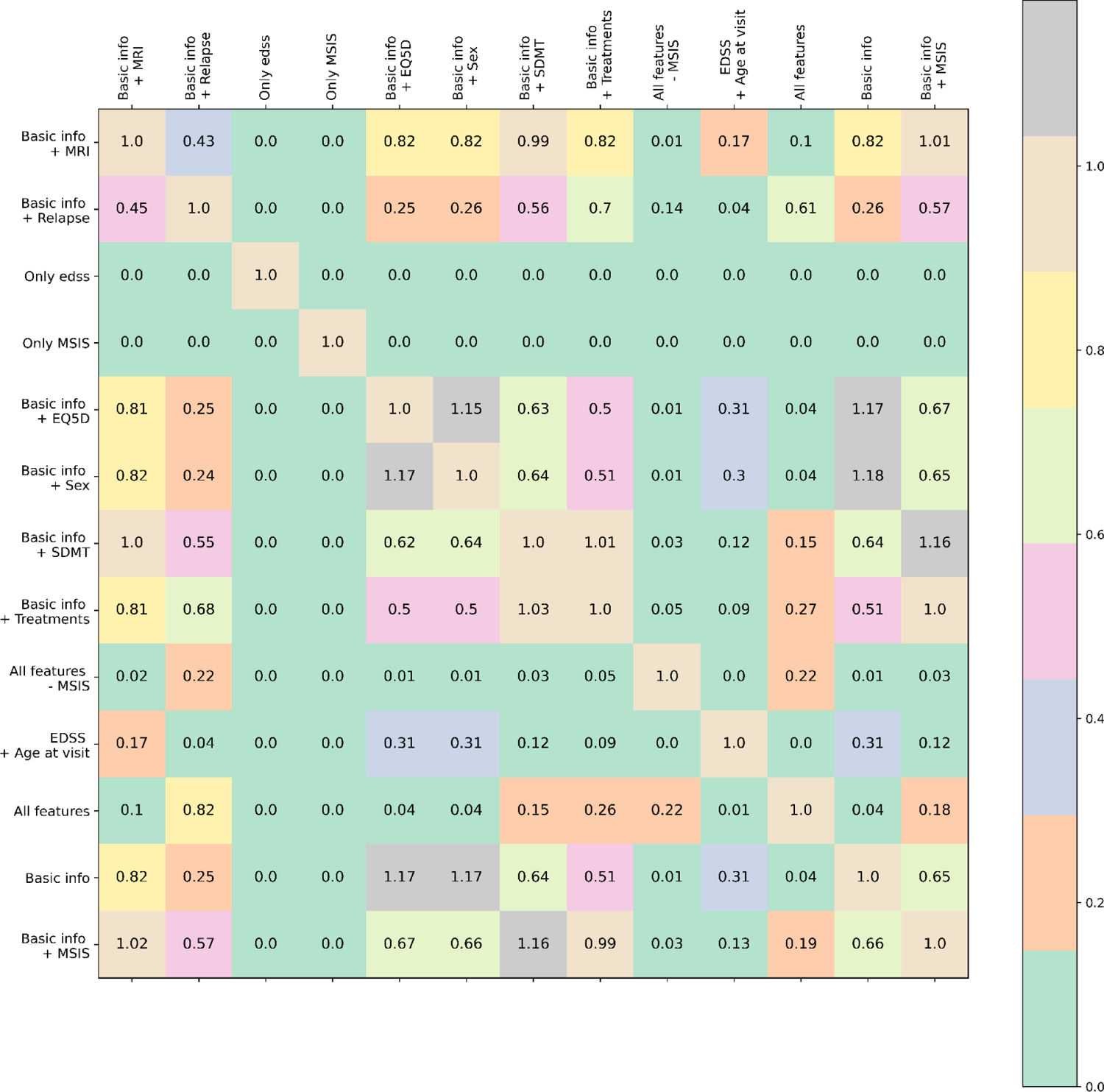
Randomization test (p-value<0.05) on F1 scores from 10-fold CV of RF models trained on combination of features. Significant p-values represent a higher or lower performance between the models. Comparing supplementary Fig. 3, the features set “All features - MSIS” has a significantly better performance than all the other combinations except “Basic info + relapse”.

**Supplementary Fig. 5:**
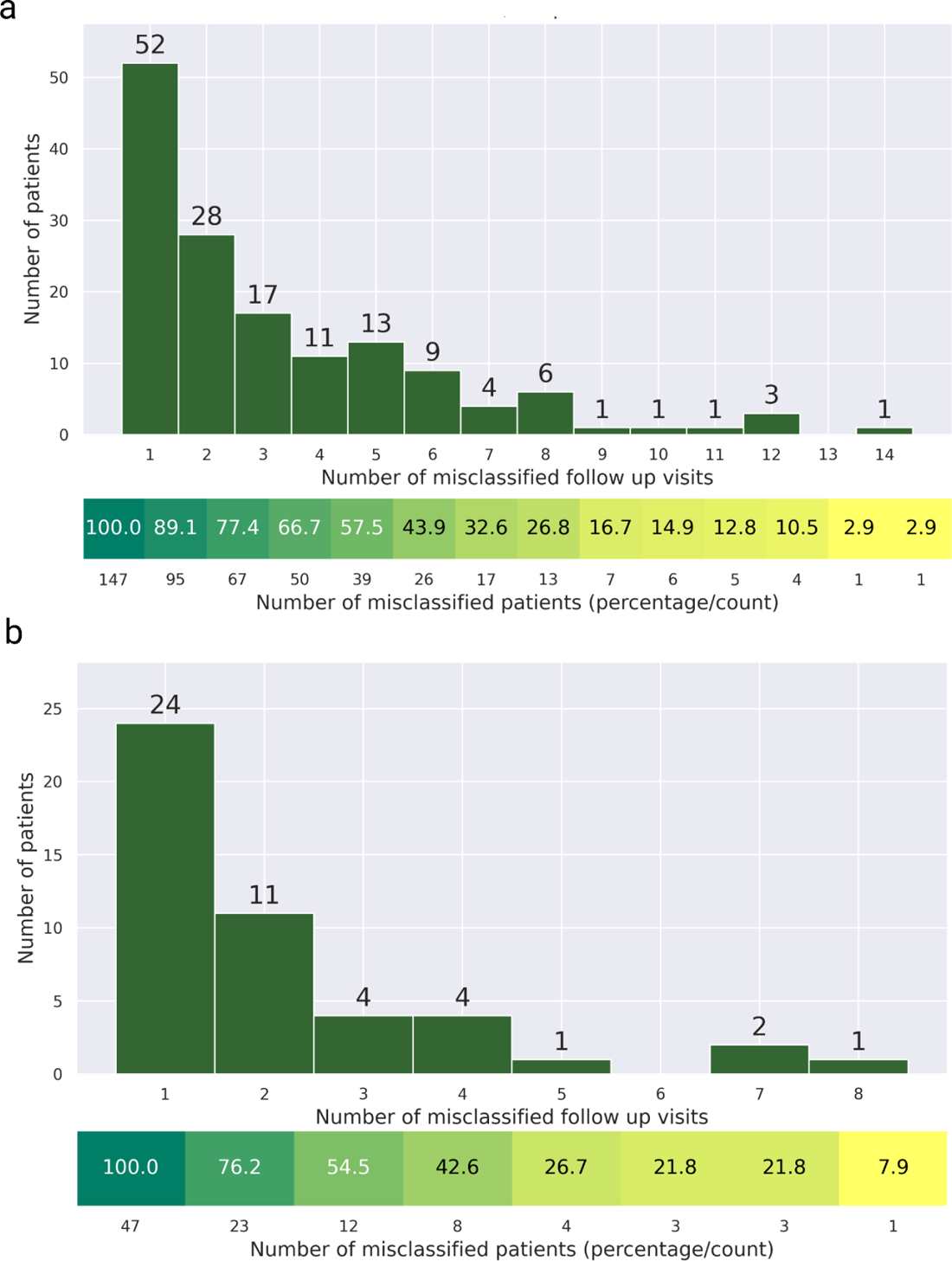
Patients and the number of misclassified hospital visits. The x-axis represents the number of misclassified hospital visits, and the y-axis represents the patients in the test set. The cumulative contribution to the erroneous prediction by patients based on the number of misclassified visits is given below. (a) The misclassification of 478 hospital visits from 147 patients diagnosed with RRMS and predicted SPMS. The 57.5% of misclassification is from 39 patients and the rest of the errors come from 108 patients. (b) The misclassification of 101 hospital visits from 47 patients diagnosed with SPMS and predicted RRMS. Twelve patients contribute to 54.5% of the misclassification, and the rest 45.5% errors come from 35 patients.

**Supplementary Fig. 6:**
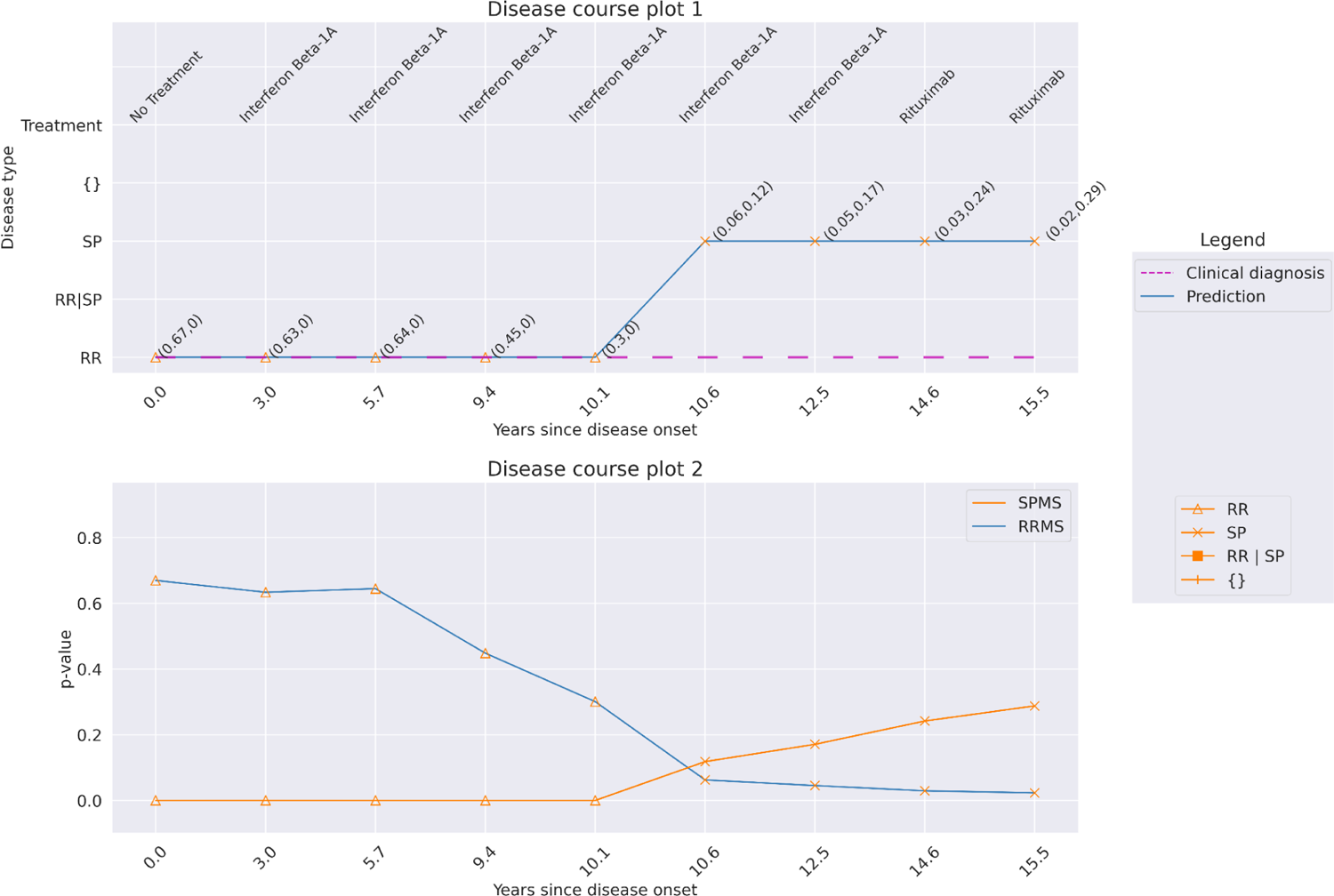
Predictions at a confidence of 92% for a patient with a disease course of 15.5 years over 9 hospital visits. The patient was diagnosed with RRMS at the onset and at the latest clinical hospital, while the prediction shows the patient has transitioned to SPMS at the hospital visit year 10.6 (Disease course plot 1). From Disease course plot 2, there is an apparent decrease in RRMS p-value between the hospital visit years 0 to 12.5, while there is an increase in SPMS p-value from the year 10.1 till the last hospital visit (15.5 years), suggesting transition from RRMS to SPMS.

**Supplementary Fig. 7:**
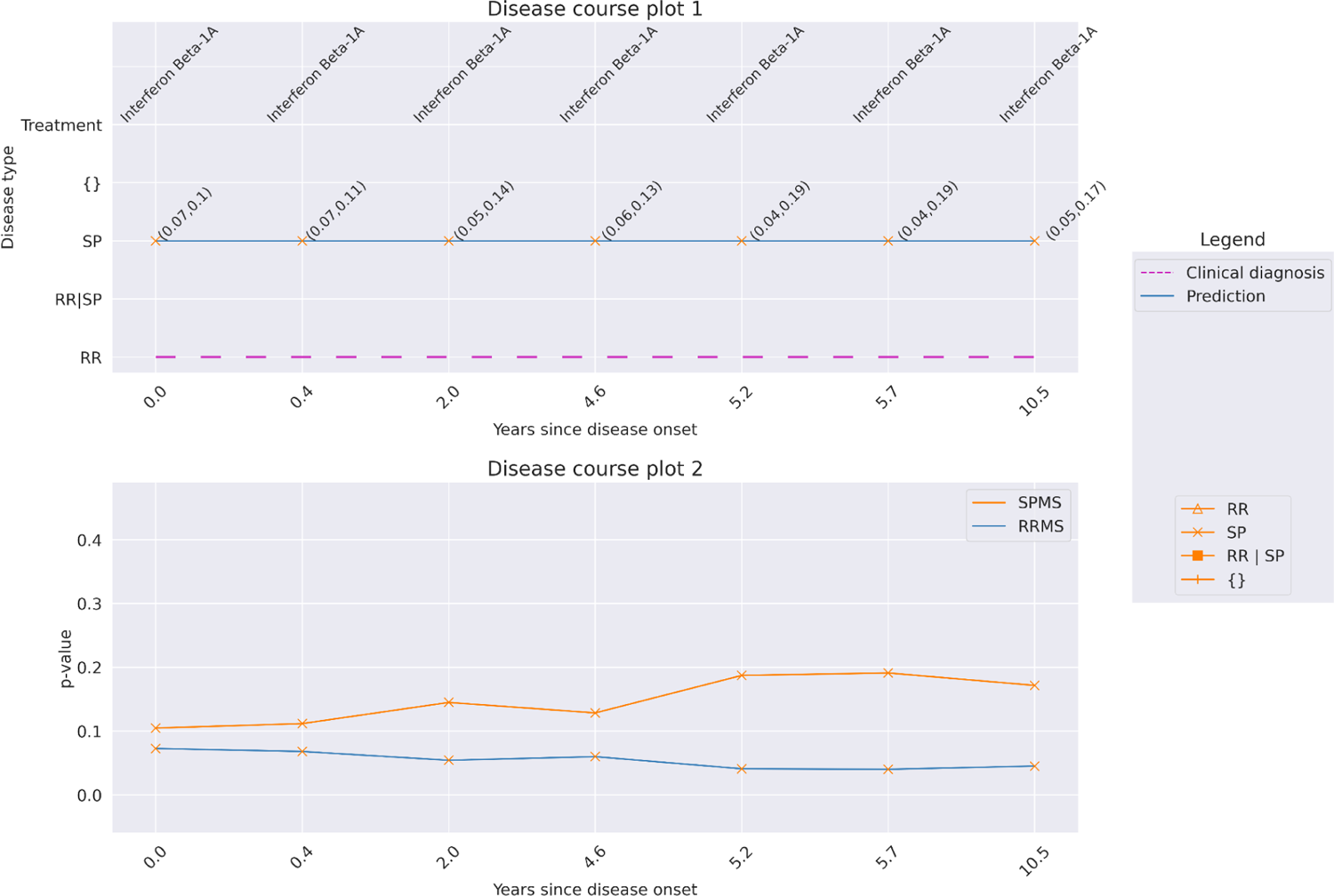
Predictions at a confidence of 92% for a patient with a disease course of 10.5 years over 7 hospital visits. The patient was diagnosed with RRMS at the onset and at the latest clinical hospital, while predicted to have SPMS from the onset (Disease course plot 1). From the Disease course plot 2, there appears to be an increased SPMS p-value compared to the RRMS p-value. Additionally, there is an increase in SPMS p-value from year 0 till year 10.5, suggesting the patient may have already transitioned to SPMS.

**Supplementary Fig. 8:**
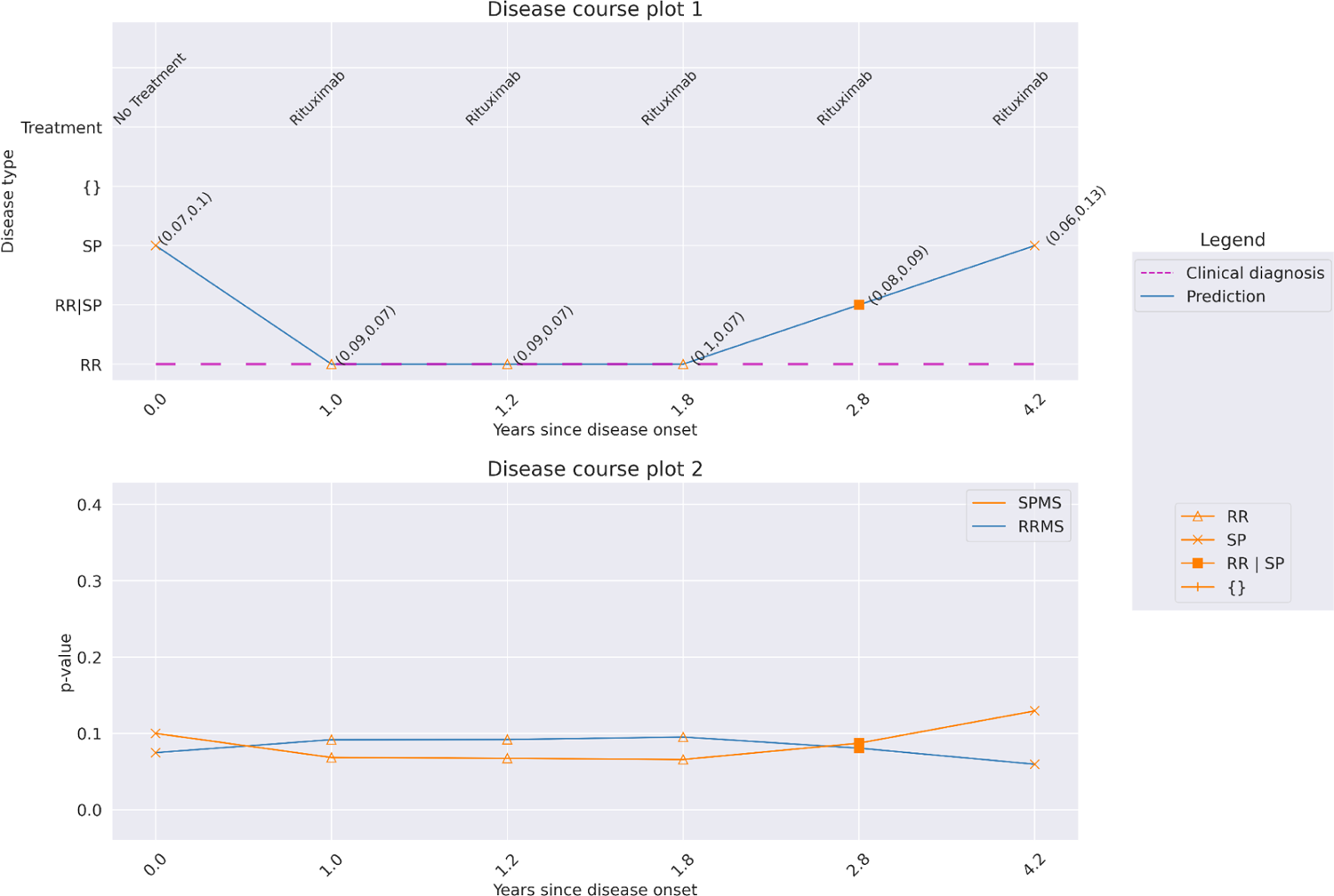
Predictions at a confidence of 92% for a patient with a disease course of 4.2 years over 6 hospital visits. The patient was diagnosed with RRMS at the onset and at the latest clinical hospital. The model makes predictions as RRMS for the visit years 1.0, 1.2, and 1.8, while the initial visit has been predicted as SPMS. The disease trajectory does not hold clinical validity, as the disease course cannot go from SPMS to RRMS. The error is likely to have been caused due to lower p-values for both RRMS and SPMS (disease course plot 2), and these errors can be minimized by increasing the confidence of the model.

**Supplementary Table 2:** Prediction made by the conformal predictor on transitioning patients. Predictions were made on 149 patients diagnosed with RRMS at onset hospital visits and SPMS at later hospital visits. The predictions can be divided into five categories based on the predictions of the initial and final hospital visits. (RRMS - SPMS means RRMS at the initial hospital visit and SPMS at the latest available hospital visit). Np=number of individual patients, nv=number of hospital visits. *Percentages would not add up due to rounding off.

| Predictions<br>(at first and latest<br>hospital visit) | Diagnosis of transitioning patients at debut and at<br>latest hospital visit. This is a subset of the SPMS<br>patients from Table 2 (np=149, nv=1775). |
| --- | --- |
| RRMS - SPMS | 107 (71.8%) |
| SPMS - SPMS | 37 (24.8%) |
| RRMS - RRMS | 3 (2%) |
| SPMS - RRMS | 1 (0.7%) |
| RRMS - Multiple-label | 1 (0.7%) |

**Supplementary Fig. 9:**
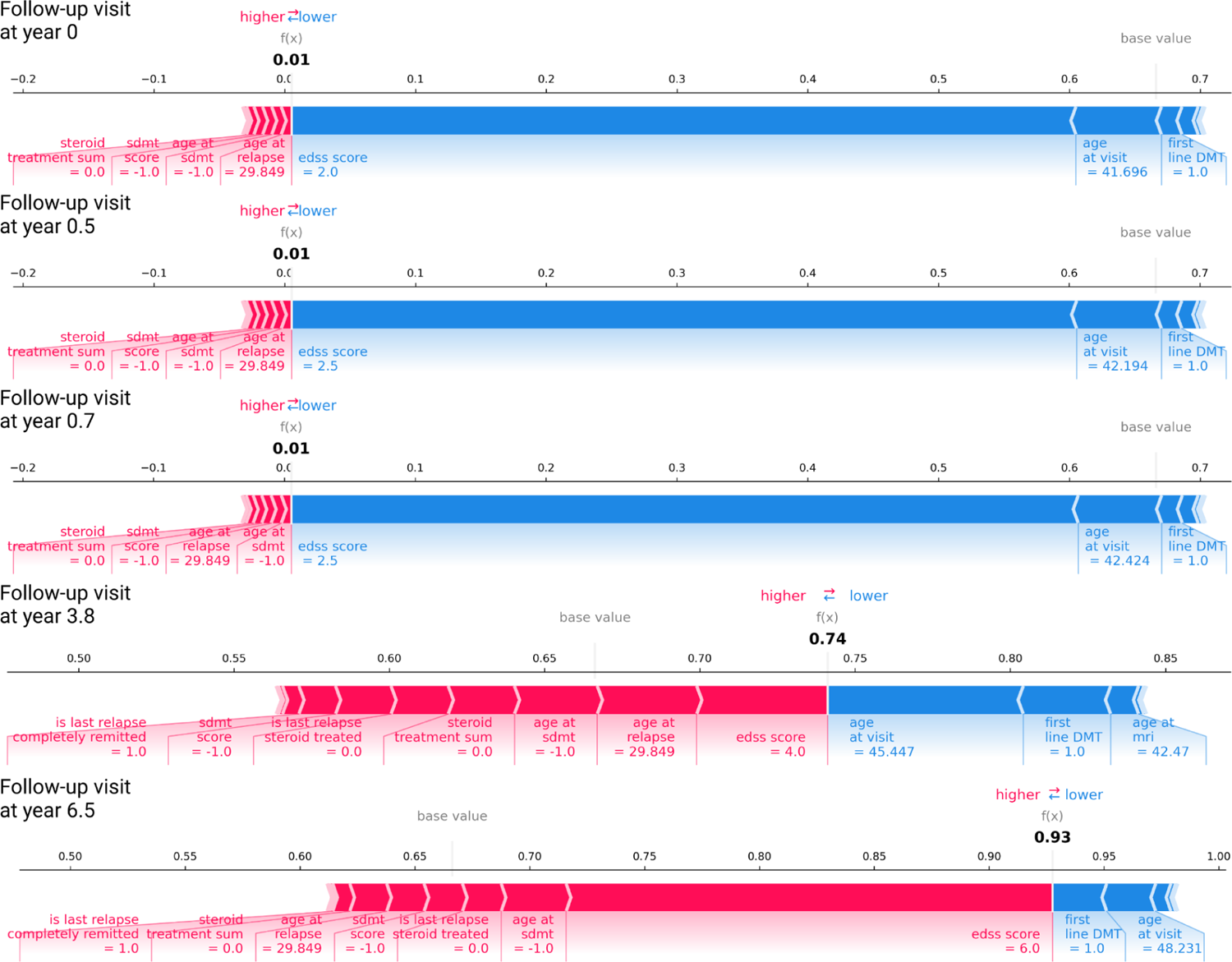
Feature contribution explanation using force plots for the predictions on the hospital visit year 0, 0.5, 0.7, 3.8, and 6.5 on the patient from Figure 3.

**Supplementary Fig. 10:**
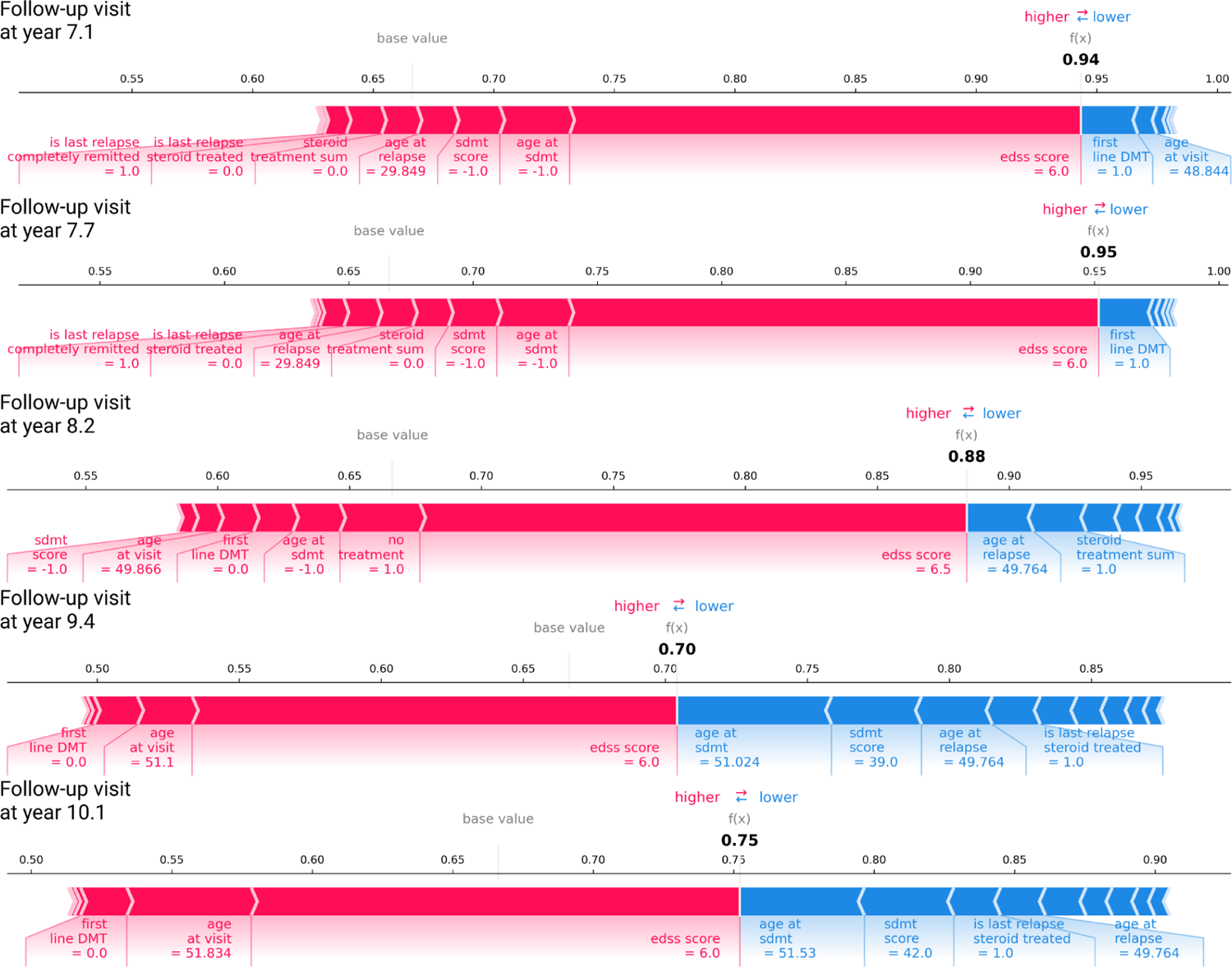
Feature contribution explanation using force plots for the predictions on the hospital visit years 7.1, 7.7, 8.2, 9.4, and 10.1 on the patient from Figure 3.

**Supplementary Fig. 11:**
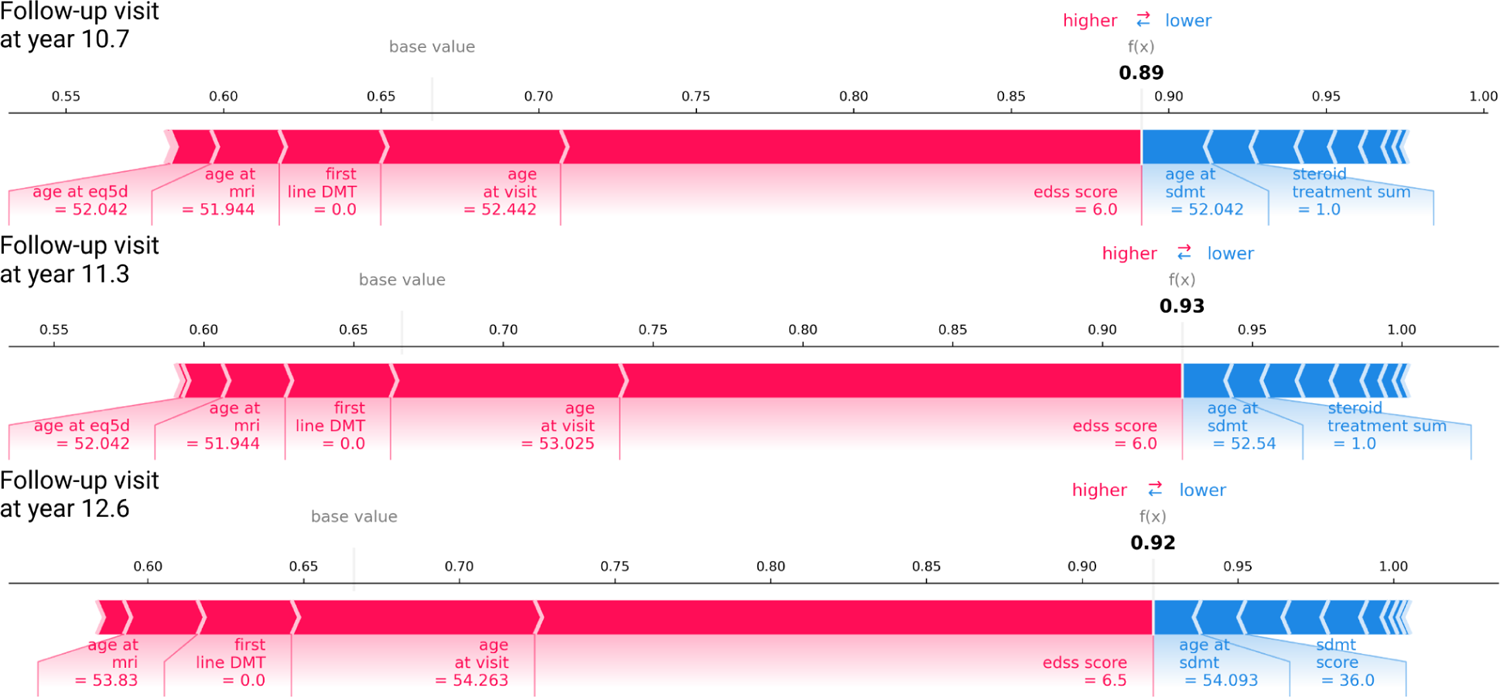
Feature contribution explanation using force plots for the predictions on the hospital visit years 10.7, 11.3, and 12.6 on the patient from Figure 3.

**Supplementary Fig. 12:**
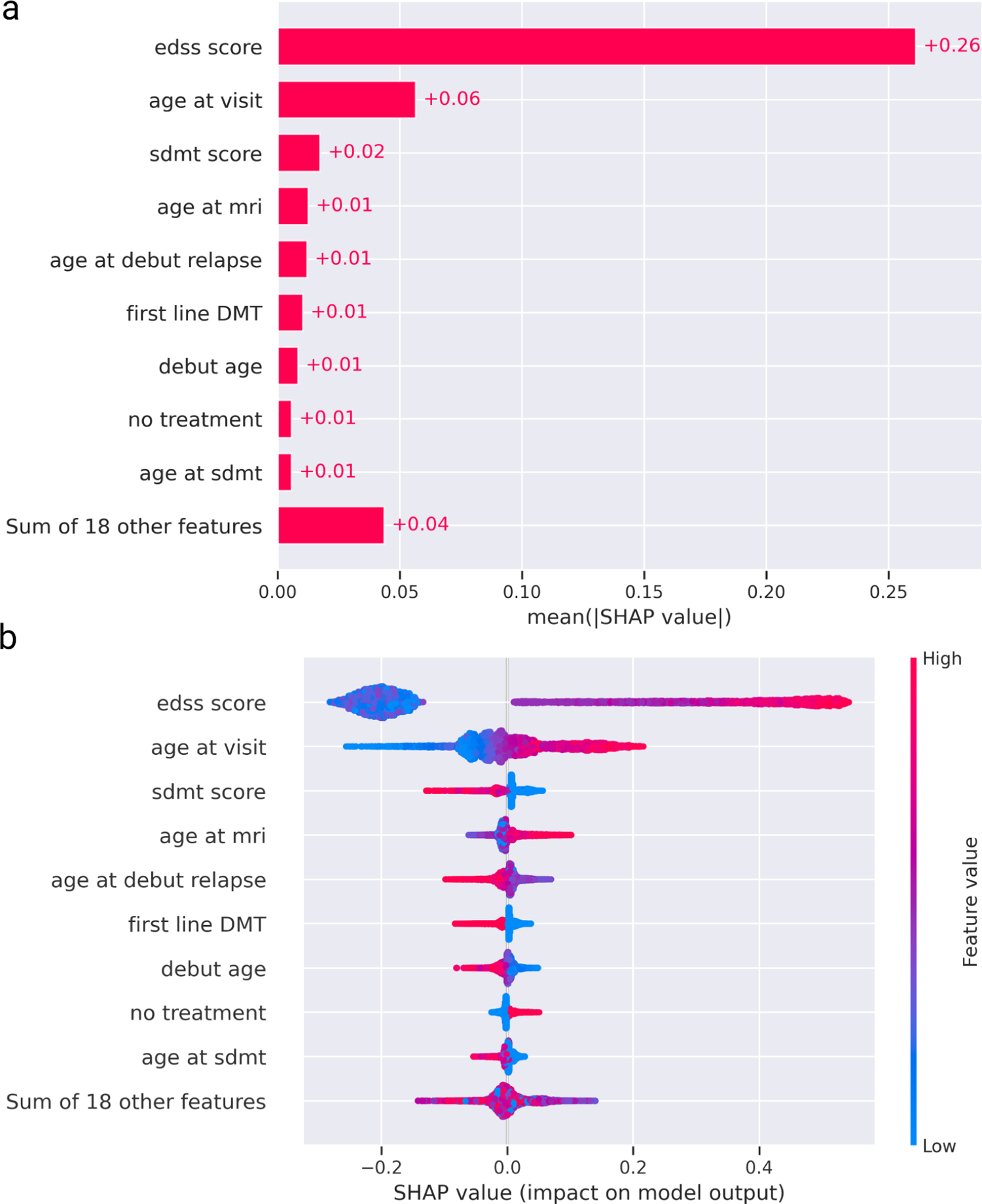
The global importance of features for prediction on the test data is explained using SHAP. (a) a summary plot using mean absolute SHAP values, (b) a bee swarm plot showing the SHAP values for each feature, calculated iteratively for each datapoint in the dataset. The values are plotted and color-coded based on their importance for each prediction.

**Supplementary Fig. 13:**
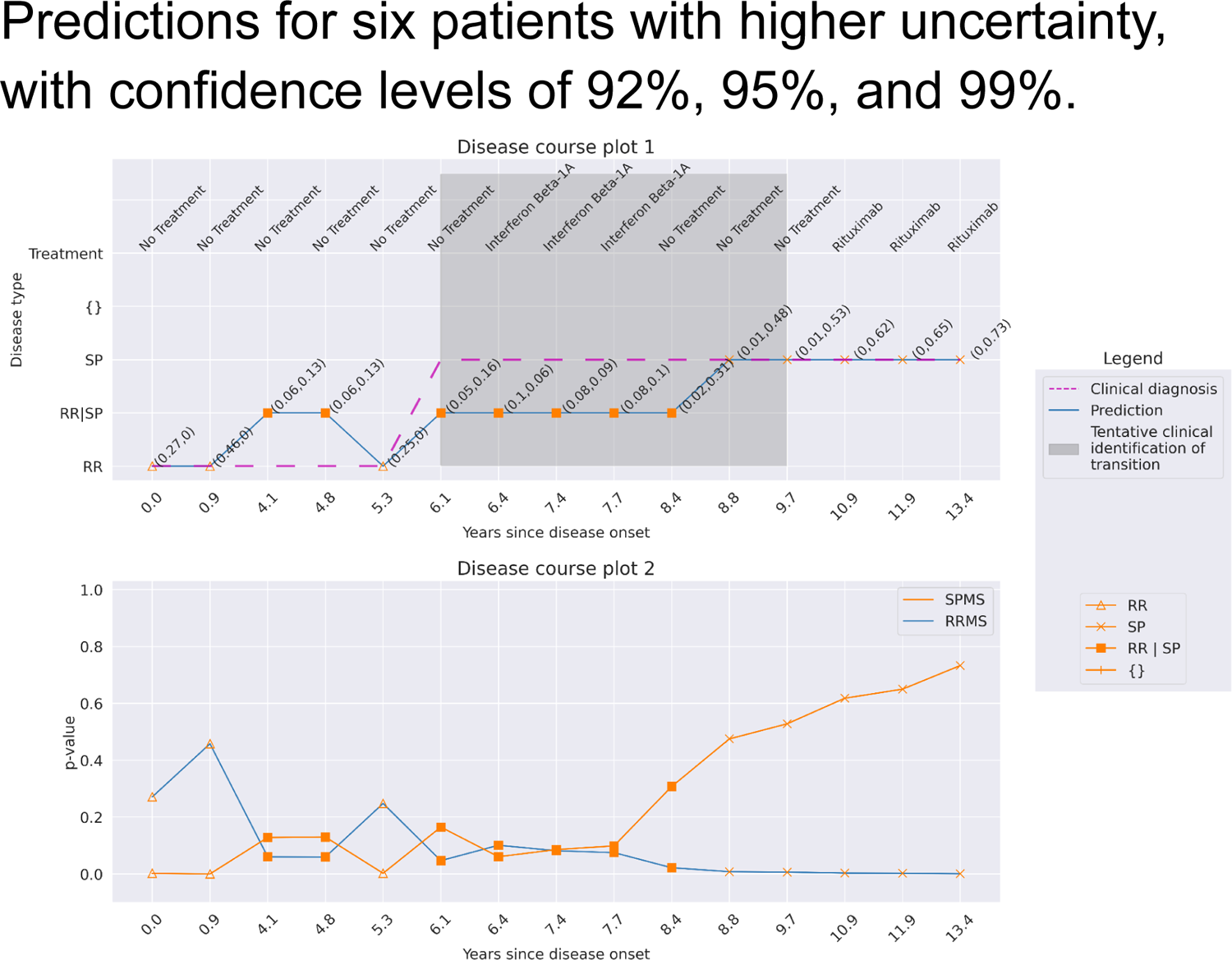
Patient 1. Predictions at a confidence of 99% for a patient with a disease course of 13.4 years over 15 hospital visits. There are more unreliable (multiple) predictions between the years 4.1 and 8.8. The predictions for years 6.1 and 8.4 became unreliable when the confidence increased from 95% to 99%.

**Supplementary Fig. 14:**
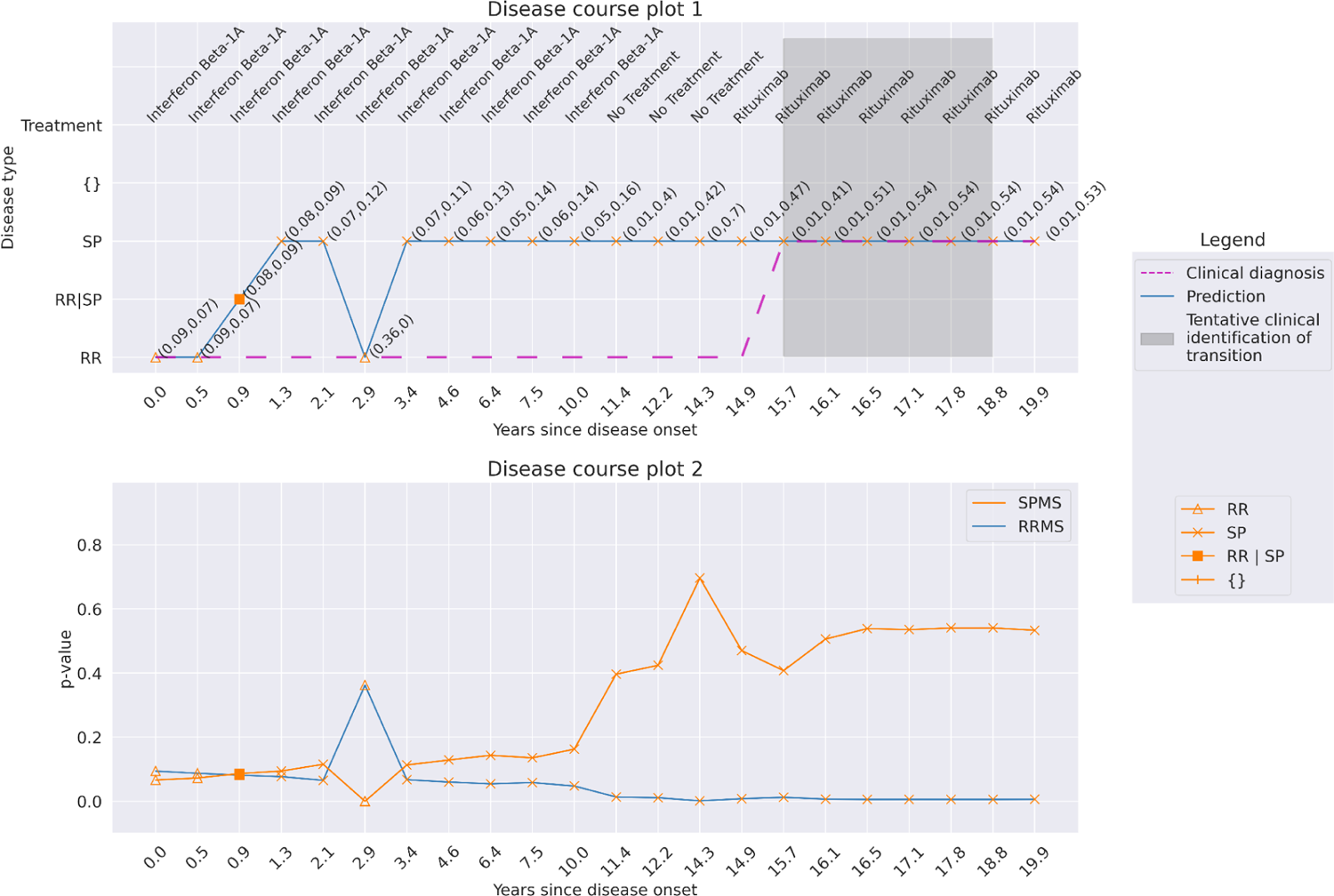
Patient 2. Predictions at a confidence of 92% for a patient with a disease course of 19.9 years over 22 hospital visits. The patient switched disease state from SPMS to RRMS at year 2.9, according to the predictions, and back to SPMS at year 3.4, which is clinically not possible.

**Supplementary Fig. 15:**
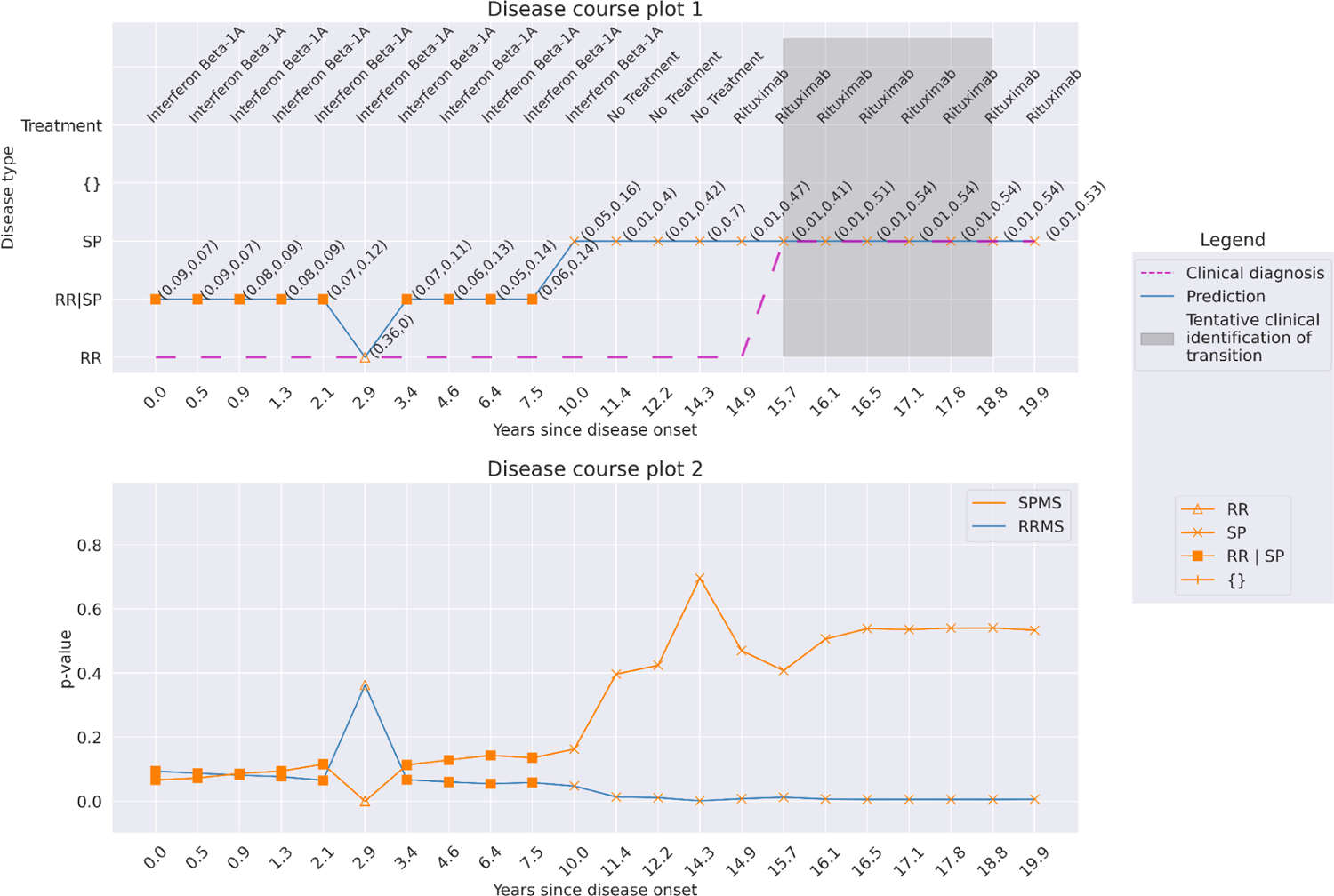
Patient 2. Predictions at a confidence of 95% for a patient with a disease course of 19.9 years over 22 hospital visits. The visits before year 2.9 and till year 7.5 became unreliable with multiple-label predictions. The disease trajectory holds clinical validity now as compared to the predictions at 92% confidence.

**Supplementary Fig. 16:**
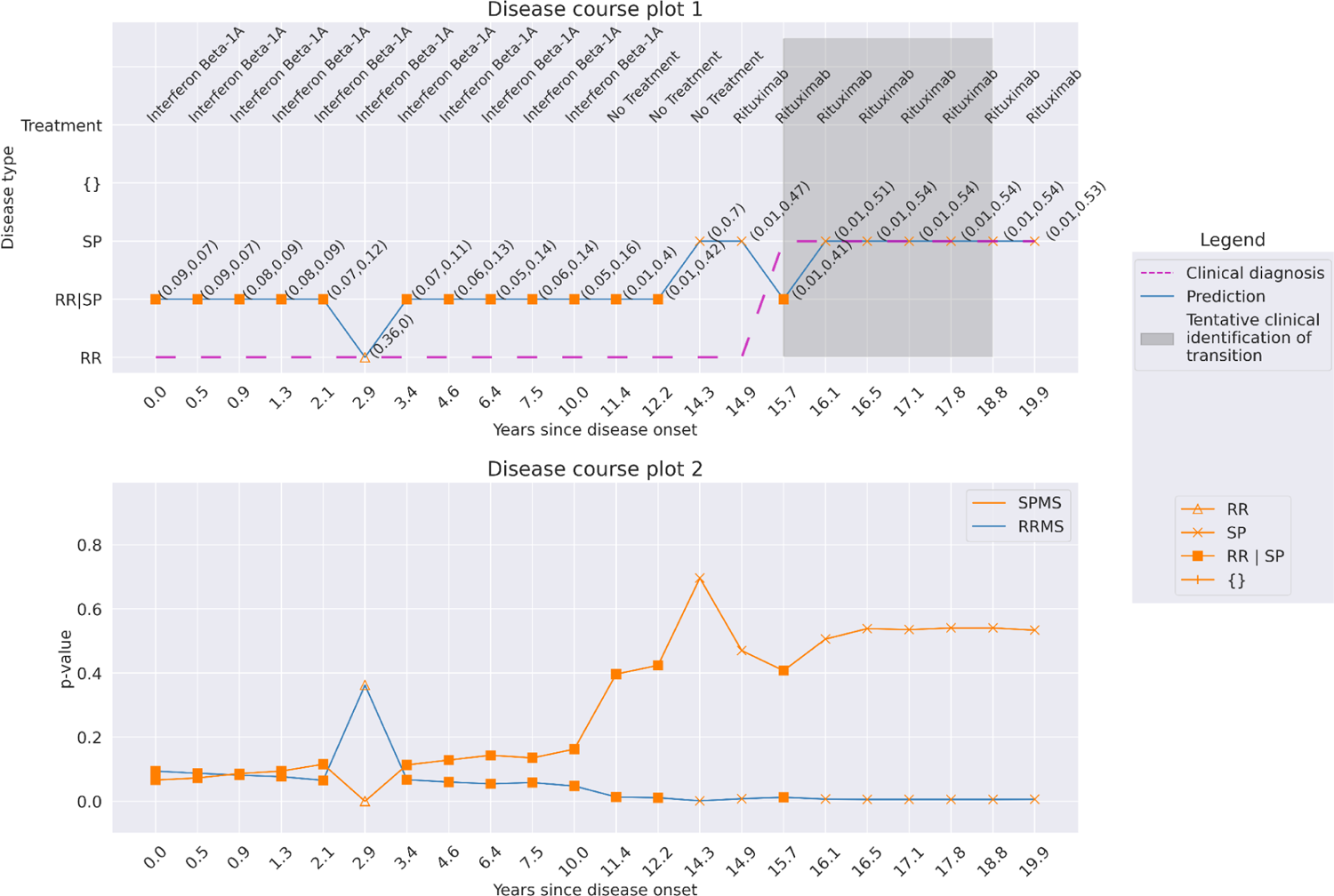
Patient 2. Predictions at a confidence of 99% for a patient with a disease course of 19.9 years over 22 hospital visits. There is an increase in multiple-label, and the predicted SPMS transition is at year 12.2, while the transition was marked at year 15.7 in the clinic.

**Supplementary Fig. 17:**
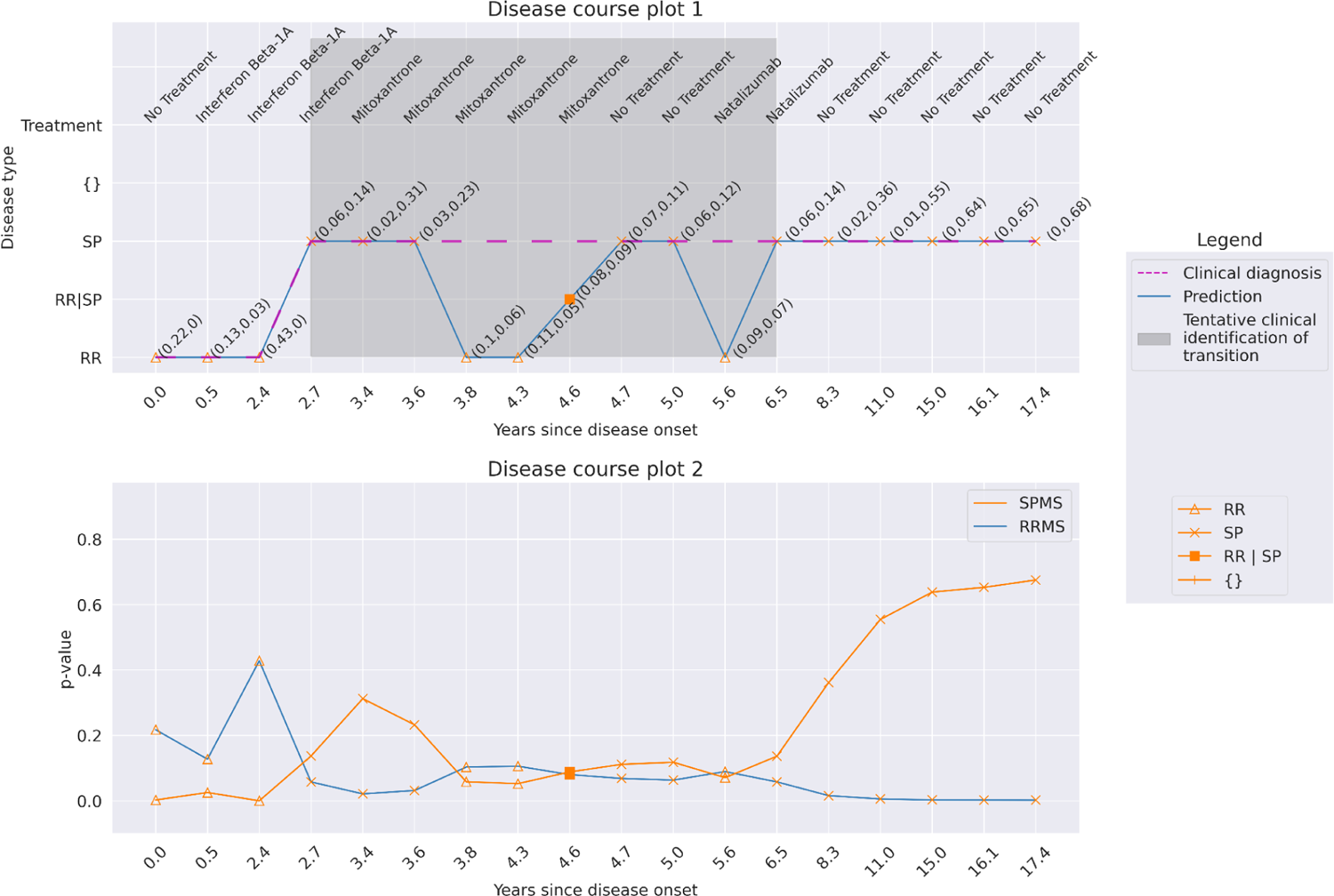
Patient 3. Predictions at a confidence of 92% for a patient with a disease course of 17.4 years over 18 hospital visits. The patient switched from SPMS to RRMS at year 3.8, while changing back to SPMS again at year 4.7. The alternating prediction can be associated with lower p-values of these predictions (Disease course plot 2).

**Supplementary Fig. 18:**
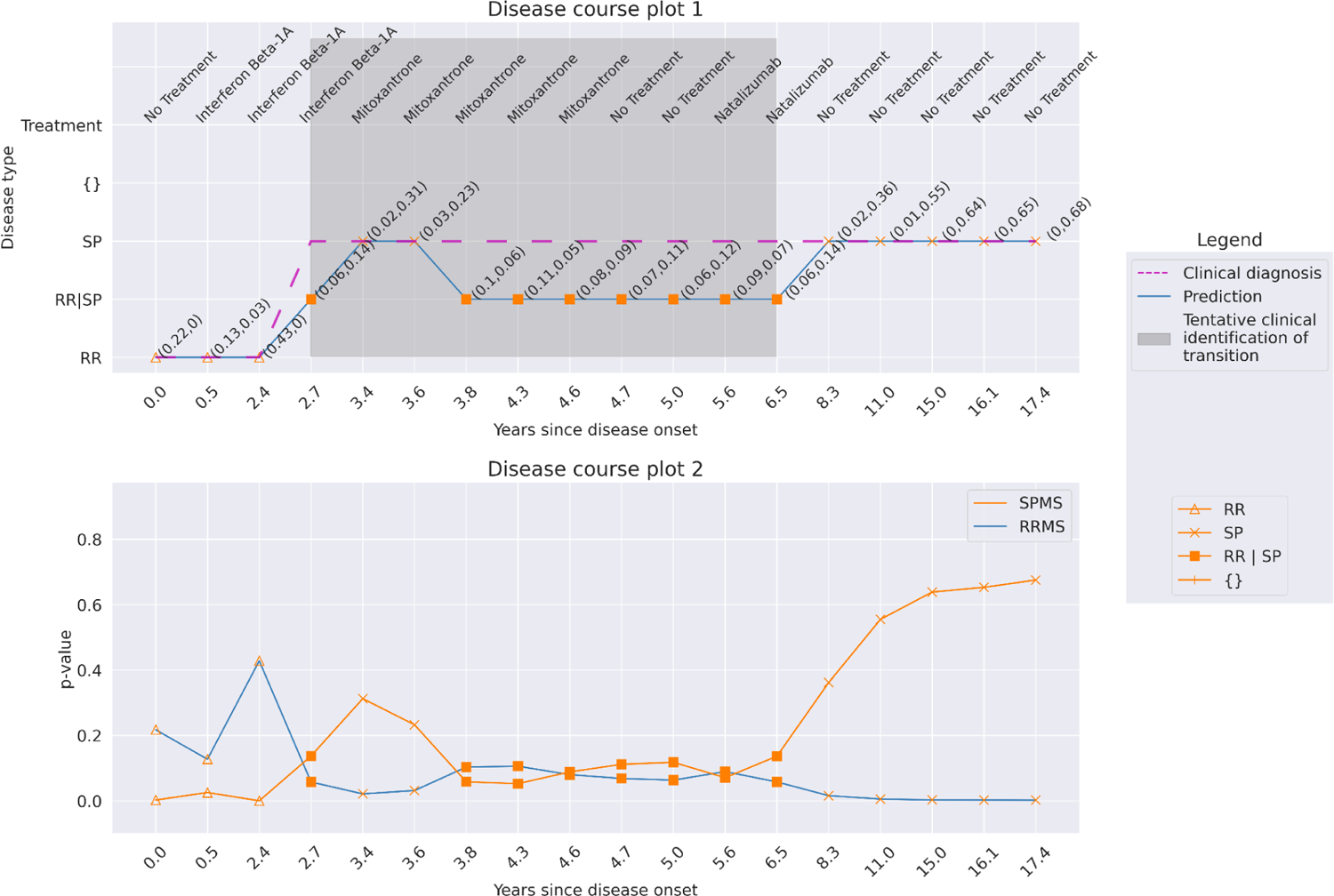
Patient 3. Predictions at a confidence of 95% for a patient with a disease course of 17.4 years over 18 hospital visits. The lower p-value predictions became multiple-label, making the disease trajectory valid. The model predicts transition at year 3.4, while clinically, it occurred at year 2.7.

**Supplementary Fig. 19:**
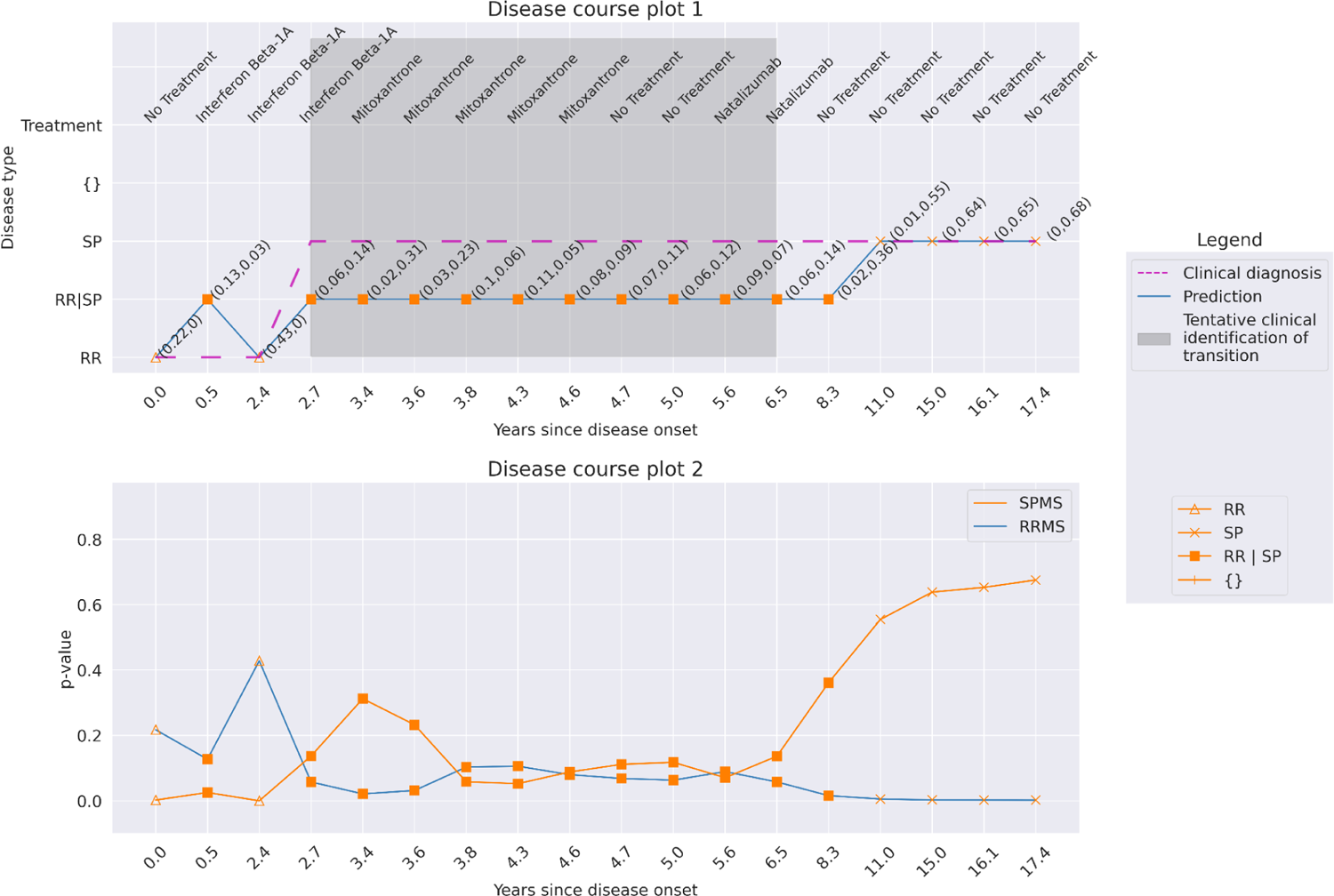
Patient 3. Predictions at a confidence of 99% for a patient with a disease course of 17.4 years over 18 hospital visits. The predictions became more stringent, marking the predictions between years 2.7 to 8.3 multiple-label. The model predicts transition at the year 11.0 at this confidence.

**Supplementary Fig. 20:**
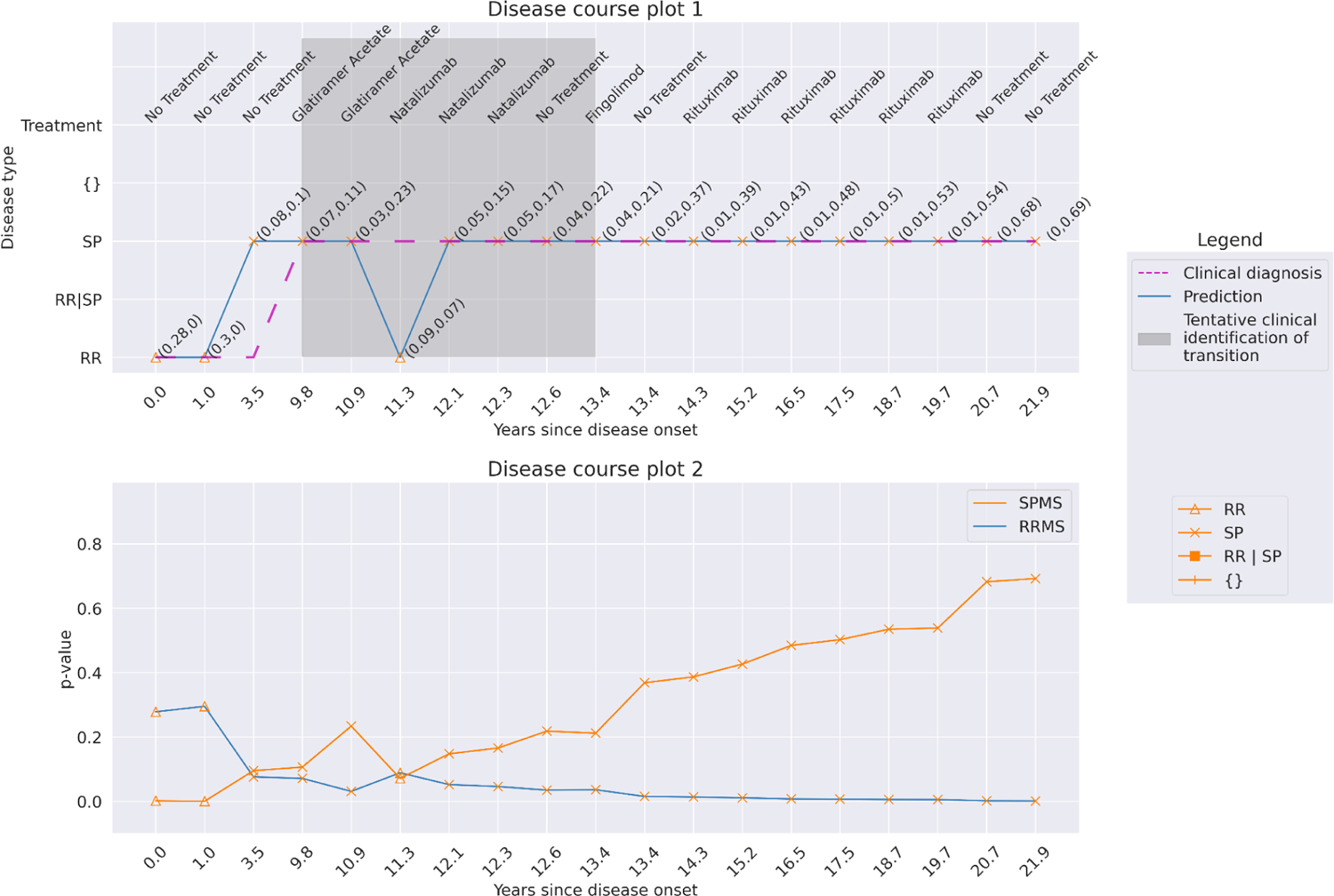
Patient 4. Predictions at a confidence of 92% for a patient with a disease course of 21.9 years over 19 hospital visits. The disease trajectory is invalid as the disease transitioned from SPMS to RRMS at visit year 11.3.

**Supplementary Fig. 21:**
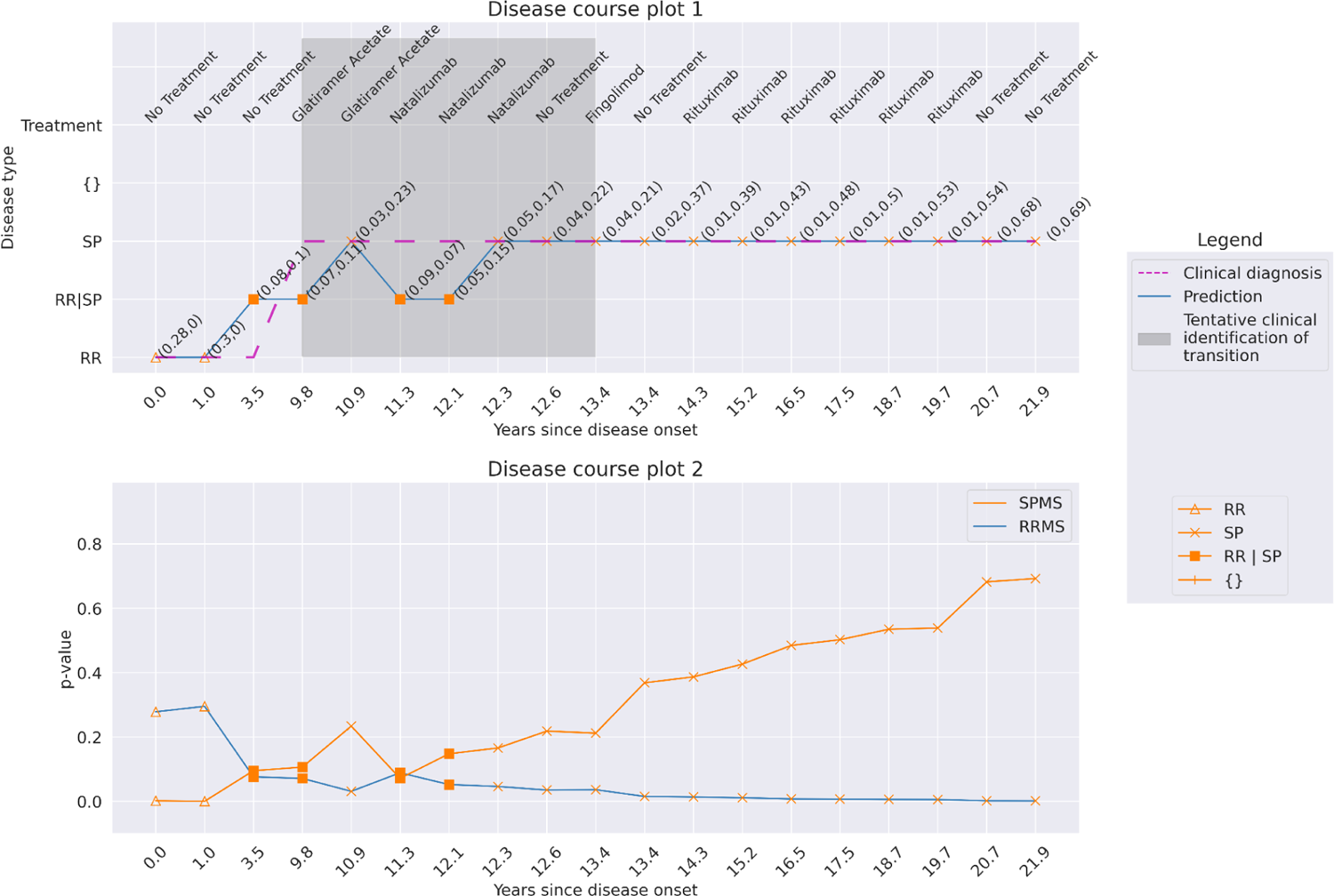
Patient 4. Predictions at a confidence of 95% for a patient with a disease course of 21.9 years over 19 hospital visits. The disease trajectory became clinically valid as the confidence increased.

**Supplementary Fig. 22:**
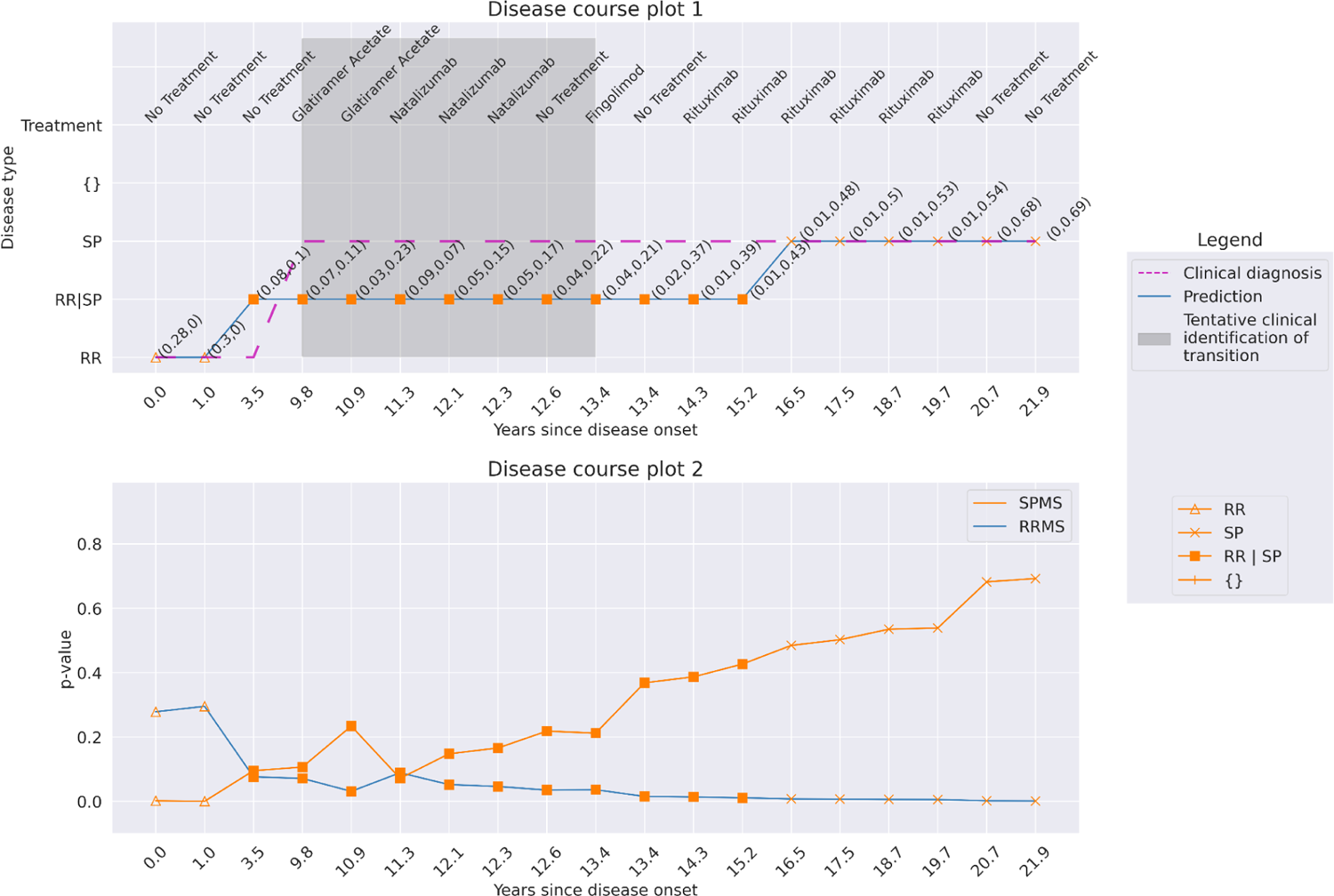
Patient 4. Predictions at a confidence of 99% for a patient with a disease course of 21.9 years over 19 hospital visits. With further increase in confidence, the model predicted transition at year 16.5 compared to year 10.9 at a confidence of 95%.

**Supplementary Fig. 23:**
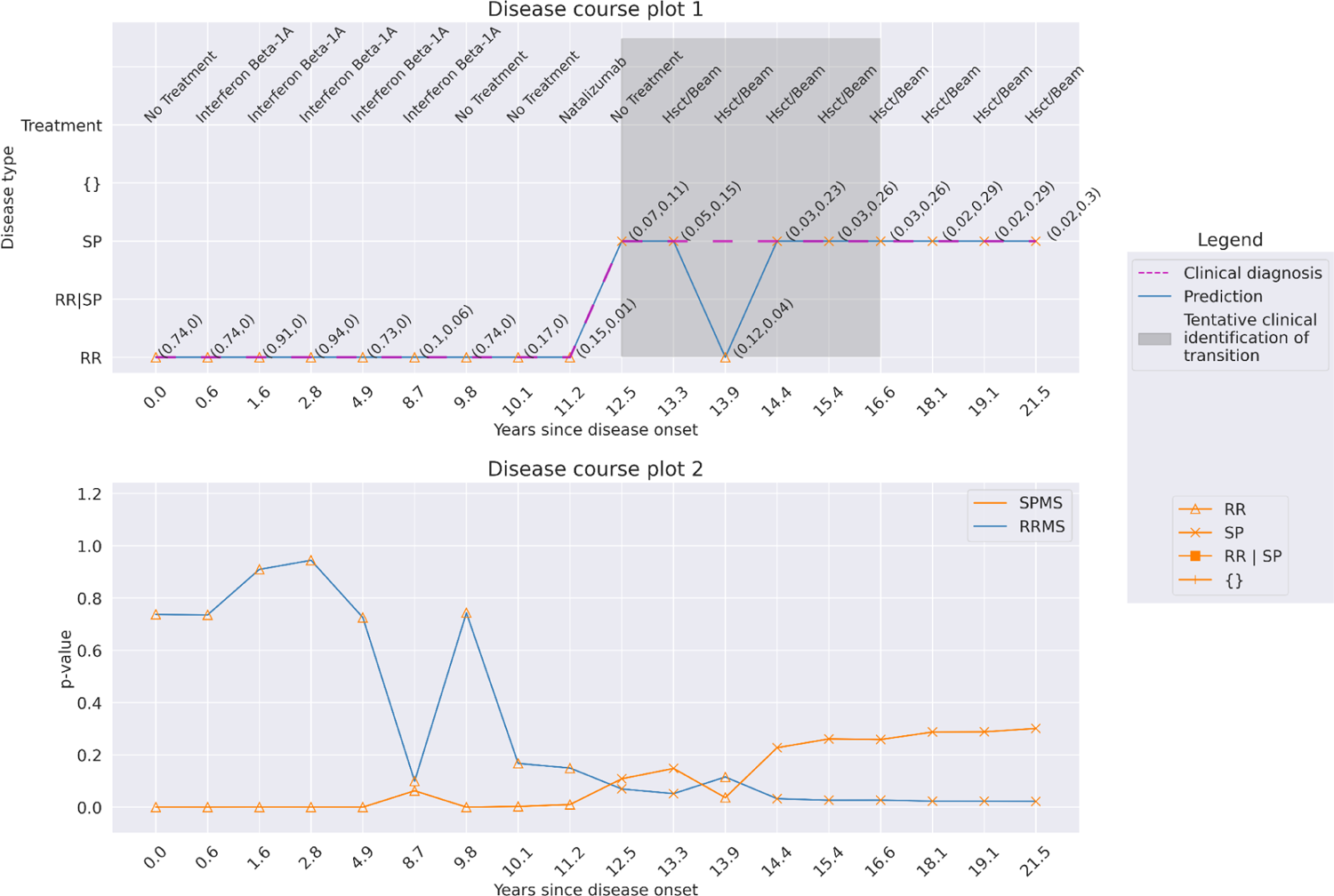
Patient 5. Predictions at a confidence of 92% for a patient with a disease course of 21.5 years and having 18 hospital visits.

**Supplementary Fig. 24:**
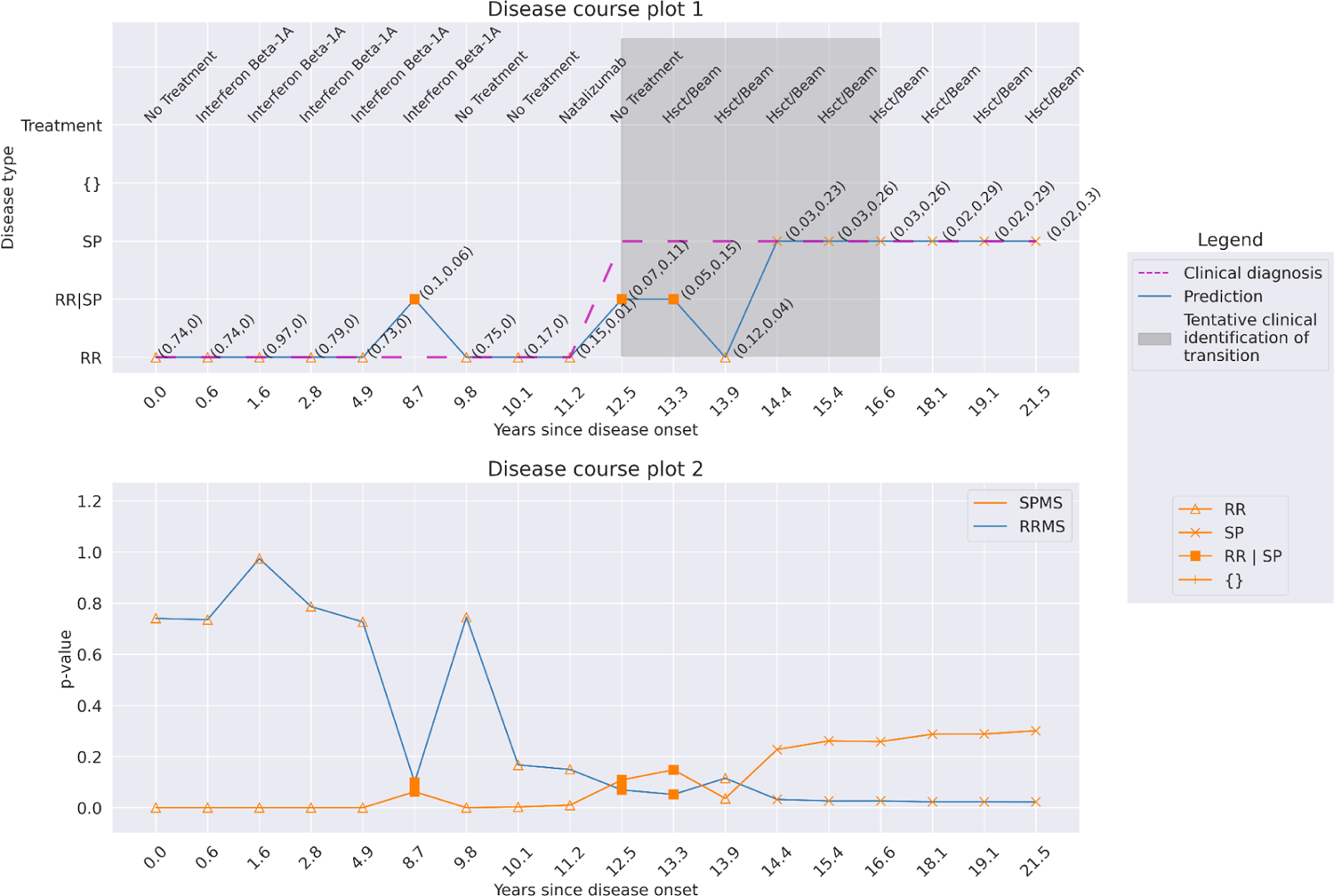
Patient 5. Predictions at a confidence of 95% for a patient with a disease course of 21.5 years over 18 hospital visits. The model produced a clinically valid disease trajectory as the confidence increased.

**Supplementary Fig. 25:**
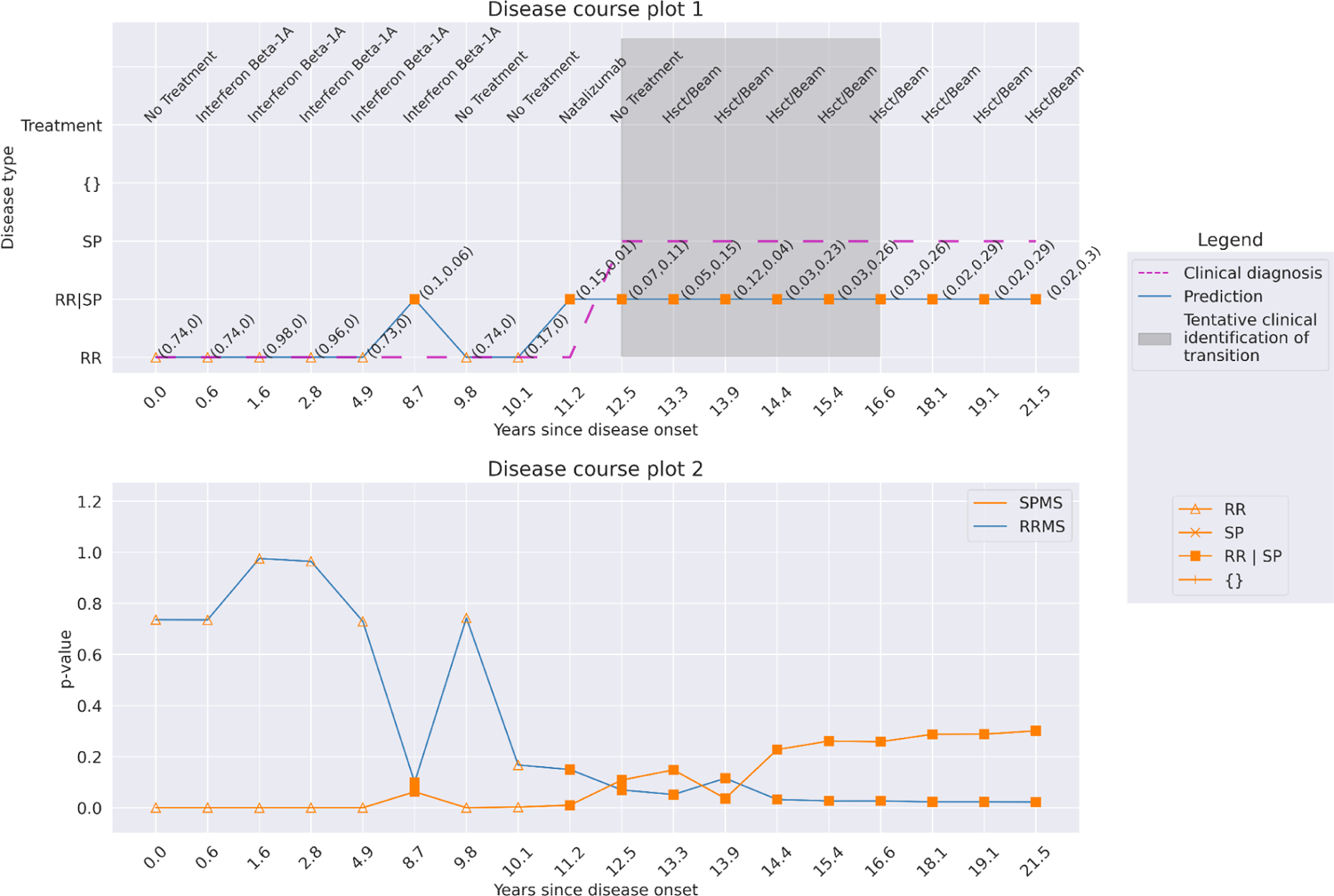
Patient 5. Predictions at a confidence of 99% for a patient with a disease course of 21.5 years over 18 hospital visits. Though there is a higher p-value for SPMS from visit 14.4 and onwards, the model does not predict single-label for these hospital visits at this high confidence.

**Supplementary Fig. 26:**
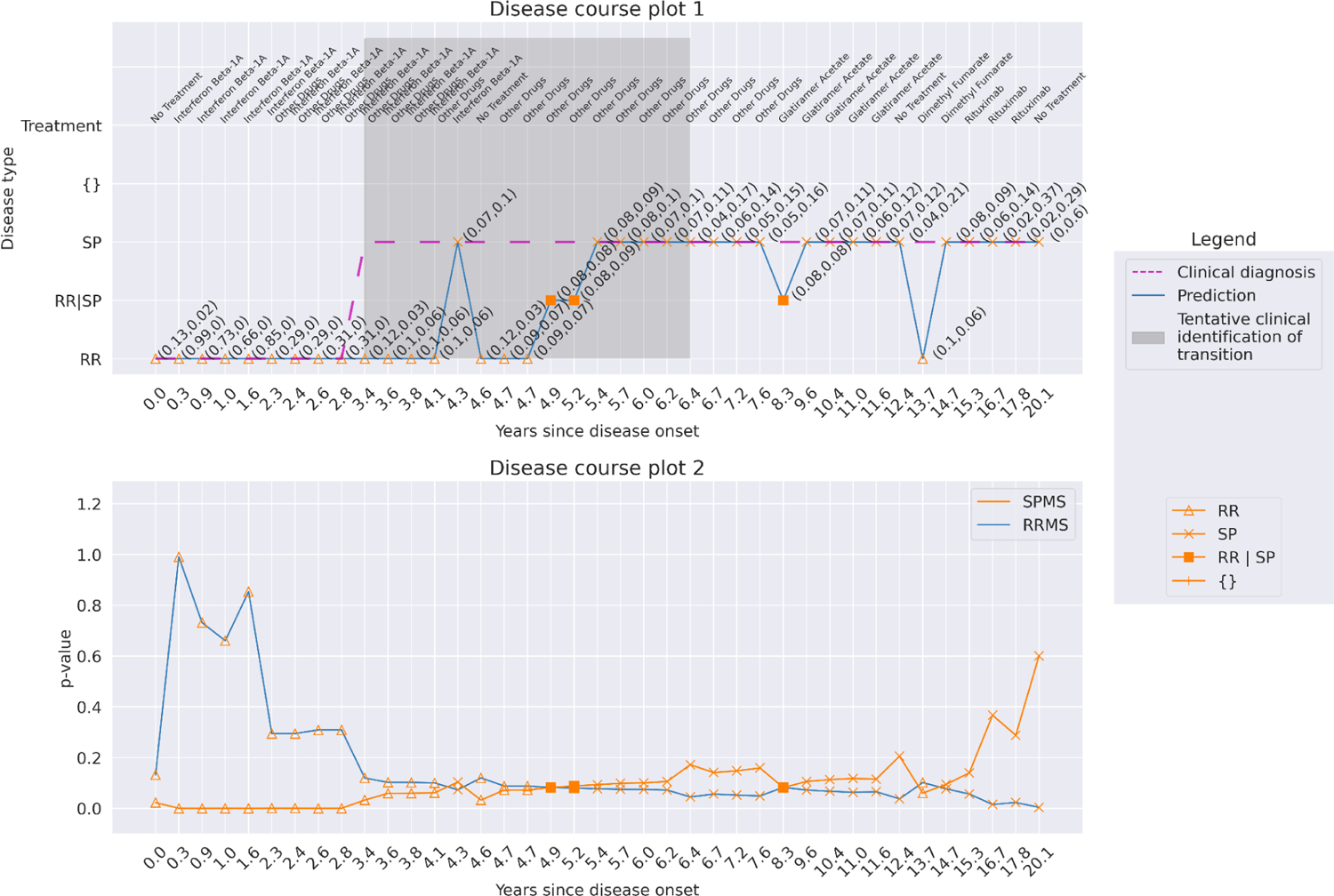
Patient 6. Predictions at a confidence of 92% for a patient with a disease course of 20.1 years over 39 hospital visits.

**Supplementary Fig. 27:**
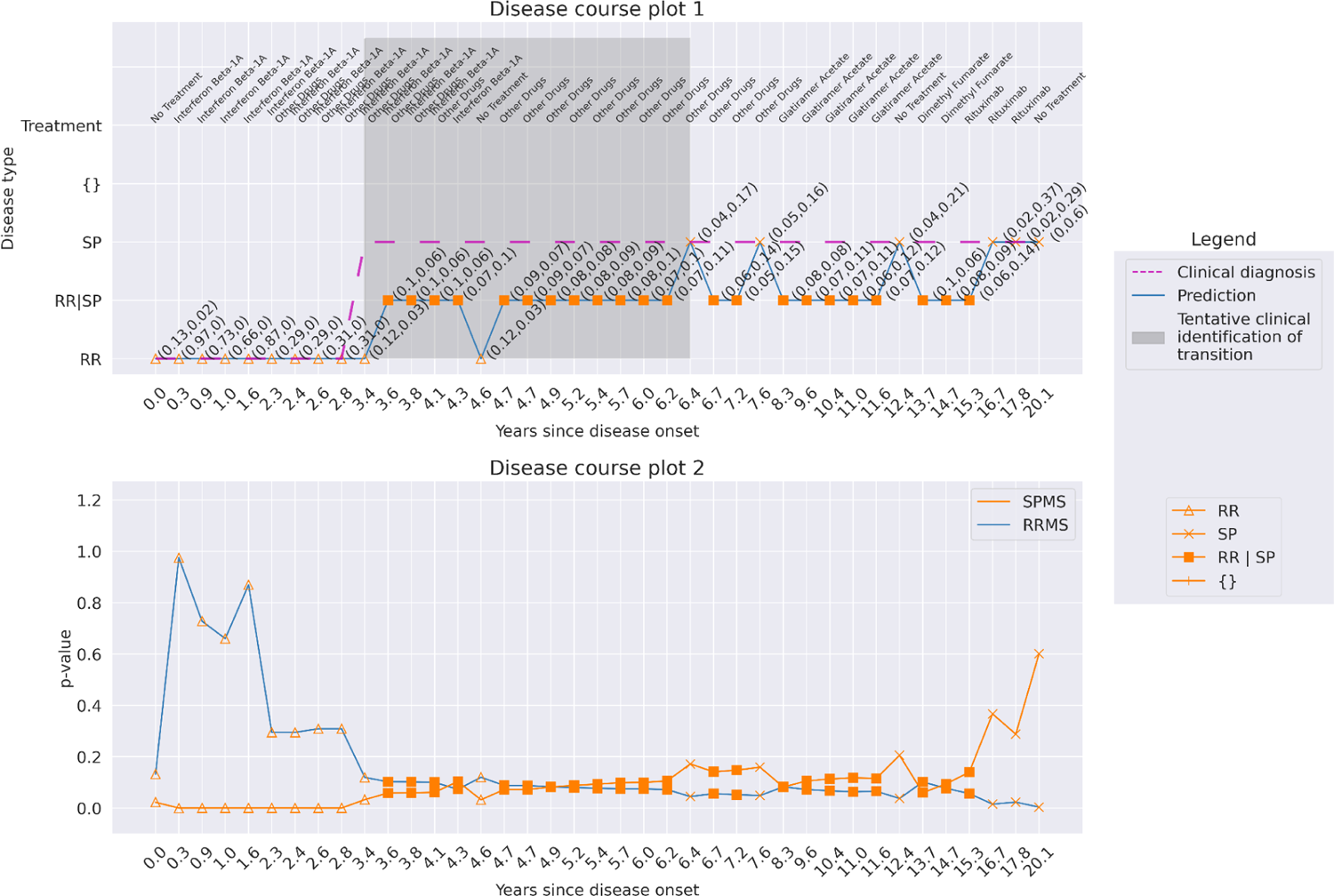
Patient 6. Predictions at a confidence of 95% for a patient with a disease course of 20.1 years over 39 hospital visits. The disease trajectory holds clinically valid, and there is an increase in multiple-label predictions. The p-values for these predictions are low, indicating the patient could be in a extended transition phase.

**Supplementary Fig. 28:**
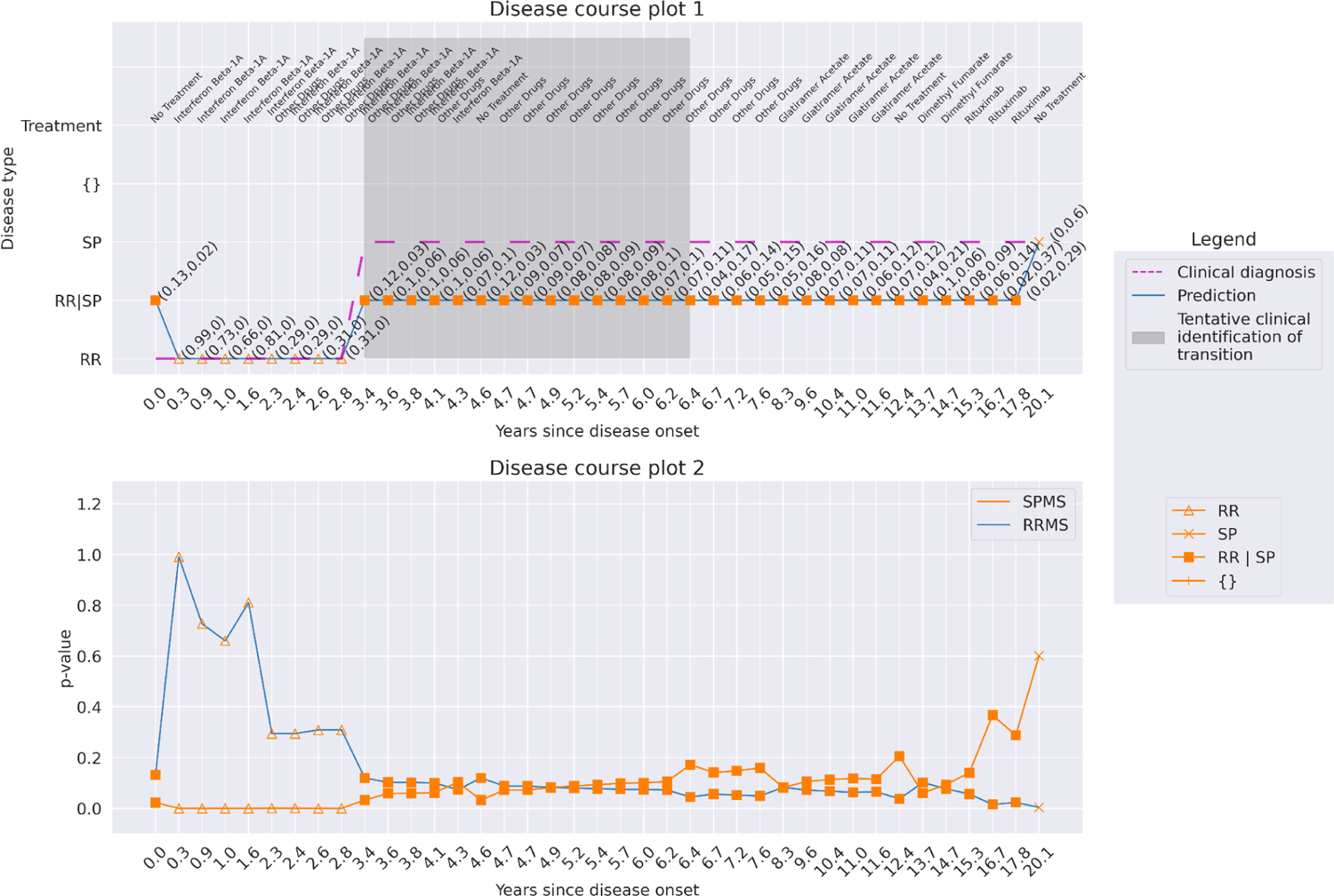
Patient 6. Predictions at a confidence of 99% for a patient with a disease course of 20.1 years over 39 hospital visits. The predictions indicate an extended transition period for a patient between year 3.4 and year 17.8.

